# Variants in *NR6A1* cause a novel oculo-vertebral-renal (OVR) syndrome

**DOI:** 10.1101/2024.11.09.24316578

**Authors:** Uma M. Neelathi, Ehsan Ullah, Aman George, Mara I. Maftei, Elangovan Boobalan, Daniel Sanchez-Mendoza, Chloe Adams, David McGaughey, Yuri V. Sergeev, Ranya AI Rawi, Amelia Naik, Chelsea Bender, Irene H. Maumenee, Michel Michaelides, Tun Giap Tan, Siying Lin, Rafael Villasmil, Delphine Blain, Robert B. Hufnagel, Gavin Arno, Rodrigo M. Young, Bin Guan, Brian P. Brooks

**Author notes:** Co-first authors (Uma M. Neelathi, Ehsan Ullah). Co-senior authors (Rodrigo M. Young, Bin Guan, Brian P. Brooks).

## Abstract

Colobomatous microphthalmia is a potentially blinding congenital ocular malformation that can present either in isolation or together with other syndromic features. Despite a strong genetic component to disease, many cases lack a molecular diagnosis. We describe a novel autosomal dominant oculo-vertebral-renal (OVR) syndrome in six independent families characterized by colobomatous microphthalmia, missing vertebrae and congenital kidney abnormalities. Genome sequencing identified six rare variants in the orphan nuclear receptor gene *NR6A1* in these families. We performed in silico, cellular and zebrafish experiments to demonstrate the *NR6A1* variants were pathogenic or likely pathogenic for OVR syndrome. Knockdown of either or both zebrafish paralogs of *NR6A1* results in abnormal eye and somite development, which was rescued by wild-type but not variant *NR6A1* mRNA. Illustrating the power of genomic ascertainment in medicine, our study establishes *NR6A1* as a critical factor in eye and vertebral development and a pleiotropic gene responsible for OVR syndrome.

## Introduction

Uveal coloboma is a congenital ocular malformation caused by failure of the ventral optic fissure to close during early eye morphogenesis and is usually considered on a phenotypic continuum with microphthalmia and anophthalmia ^1–5^. A rare condition^6–11^, coloboma may nonetheless account for up to 10% of childhood blindness^12^. Although significant progress has been made in identifying genes associated with syndromic and non-syndromic coloboma, the yield of diagnostic testing remains low, especially for isolated, non-syndromic coloboma, suggesting other genes are yet to be discovered^13–15^. To identify novel coloboma genes, the National Eye Institute has conducted a natural history study since 2006, on the genetics of coloboma that includes systematic deep phenotyping of probands and first-degree family members. We have previously identified a novel syndrome characterized by missing vertebrae (in the thoracic and/or lumbar spine) and uveal coloboma, inherited in an autosomal dominant fashion with incomplete penetrance and variable expressivity^16^.

We identified structural and sequence variants in the transcription factor gene *NR6A1* (*Nuclear receptor subfamily 6, group A, member 1,* OMIM*602778) in three families by genome sequencing (GS). These results were extended via analysis of the Genomics England 100,000 Genomes Project (UK100KGP), where three additional individuals with microphthalmia/anophthalmia/coloboma were identified ^17^.

Originally termed germ cell nuclear factor (*GCNF*)/retinoid receptor-related testis-associated receptor (*RTR*), *NR6A1* is an orphan member of the nuclear hormone receptor family of transcription factors, often acting as a transcriptional repressor. *NR6A1* is highly expressed in embryonic and other stem cells from various tissues (especially testes) and is repressed upon differentiation^18^. *NR6A1* plays an important role in somite and subsequent vertebral development in mice, and in livestock species it is correlated with vertebral number ^19–22^. To our knowledge there are no reports on the role of *NR6A1* in eye or kidney development.

Here we described a novel autosomal dominant oculo-vertebral-renal (OVR) syndrome caused by variants in the orphan nuclear receptor gene *NR6A1*, supporting the pathogenicity of variants through a combination of in silico, in vitro and in vivo investigations. To our knowledge, this is first mendelian trait in humans characterized by missing vertebrae.

## Methods

### Patients and clinical studies

Complete eye examinations and genetic testing at the National Eye Institute (NEI) were conducted under IRB-approved clinical protocols (NCT01778543, NCT01087320, NCT02077894, www.clinicaltrials.gov). Probands underwent systemic testing as clinically indicated, which include physical exam, kidney ultrasound, routine blood chemistries, audiology, and spine x-ray. Eye examinations included age-appropriate testing of visual acuity, refraction, ocular motility/alignment, slit lamp exam, dilated fundus exam and ophthalmic photography. Specific informed consent for exome/genome sequencing was obtained under an IRB-approved protocol along with pre- and post-test genetic counseling (NCT02077894). Family COL005 and COL034 were previously reported as Family 1 and 2, respectively, without molecular characterization and detailed individual phenotyping data^16^. For patients and relatives recruited from the Genomics England 100,000 Genomes Project (UK100KGP), informed consent for whole genome sequencing (GS) was obtained in accordance with approval from the HRA committee East of England-Cambridge south (REC 14/EE/1112)^17^. Details of gene/protein expression, bioinformatic, molecular modeling, and zebrafish experiments are described in detail in Supplemental Materials and Methods.

## Results

### Variants in NR6A1 cause an oculo-vertebral-renal (OVR) syndrome

We identified three rare *NR6A1* variants in three families affected by uveal coloboma (COL005, COL034, COL171) with or without microphthalmia, cataract, and missing vertebrae through genome sequencing. In cases where multiple generations are affected, transmission is autosomal dominant with incomplete penetrance and variable expressivity (Fig. 1A). (Note that, in compliance with MedRx policy regarding potentially identifying information, all panels of Figure 1 have been deleted from the preprint server but are available upon request from the corresponding author). Clinical data for all the participants with a positive molecular result in shown Table 1. No other candidate pathogenic variants in *NR6A1* were identified in the NEI coloboma cohort consisting of a total of 224 probands (66 analyzed by genome sequencing, 57 by exome sequencing, and 101 by amplicon sequencing).

**Fig. 1:** Phenotypes associated with variants in *NR6A1*. **A**. Pedigrees of three families (COL005; COL034; COL171) from the NEI cohort demonstrating coloboma with or without microphthalmia and cataract, missing vertebrae, and congenital renal anomalies. Inheritance is autosomal dominant with incomplete penetrance and variable expressivity. **B**. Linear pigmentary disturbance representing a *forme fruste* of coloboma (arrow) in COL005.1 (right eye). **C**. Larger chorioretinal coloboma in the left eye of COL005.1 demonstrating a retinal tear in the far periphery (arrowhead). **D**. Iris coloboma of the left eye of COL005.10. **E.** Microphthalmia of the left eye in COL034.1. **F**. Retroillumination image of the left eye of COL171.1 demonstrating iris coloboma and posterior subcapsular cataract (open arrow). **G**. Spine x-ray of COL005.4 demonstrating 11 thoracic (normal 12) and 4 lumbar (normal 5) vertebrae. **H**. Schematic of NR6A1 variants detected in the NEI and UK Genomics England cohorts. +, individual with variant; -, individual without variant. DNA binding domain (DBD) and putative nuclear receptor ligand binding domain (NR-LBD) are noted. **Pedigrees of the Probands and clinical images of eyes and spine are available upon request from the corresponding author.**

The proband of the family (COL005.1) was presented with bilateral uveal colobomas (Fig. 1B, C). Family history was notable for a sibling (COL005.4), and relatives (COL005.10), (COL005.17) with uveal coloboma. The deletion breakpoints were in intron 2 and 6 removing the coding sequence for amino acids (aa) Ile48-Gly275 and likely causing a frameshift (p.Ile48Asnfs*3, Fig. 1H). The status of the heterozygous deletion was determined by breakpoint PCR among family members available, which revealed complete segregation with the missing vertebrae with an estimated LOD score of 3.6 (Fig. 1A, Supplementary Fig. S1). Four family members were also affected by coloboma in addition to missing vertebra, of which one (COL005.17) also had, by report, only one kidney. The proband of family (COL034.1) was presented with bilateral uveal colobomas and microphthalmia OS (Fig. 1E). Genome sequencing revealed a heterozygous c. 274C>T p. (Arg92Trp) variant in *NR6A1*, which was found in the affected parent and essentially unaffected grandparent (Fig. 1A, H). The proband of family (COL171.1) was presented with bilateral colobomatous microphthalmia affecting the iris, retina/choroid and optic nerve. Slit lamp exam was notable for bilateral microcornea, bilateral posterior subcapsular and nuclear cataracts and missing zonules inferiorly OU (Fig. 1F). Genome sequencing revealed a heterozygous c.1306C>T p.(Arg436Cys) variant in the proband which was absent in his unaffected parent (Fig. 1H). Detailed study of the probands and their family members available for evaluation, were described in clinical vignettes in the extended data. No convincing pathogenic variants in known coloboma genes were identified in any of these subjects.

### Genome-first approach for NR6A1 variants corroborates MAC phenotypes

We performed an unbiased disease association analysis of rare pLoF variants using the UK100KGP dataset^17^. After removing variants resulting from calling artifacts or mis-annotation, only three pLoF variants were found in the cohort with approximately 126,700 alleles (Supplementary Table 4, Supplementary Fig. 2). We found three probands, Proband (A1) with bilateral chorioretinal coloboma (*forme fruste* OD) and OS coloboma of the optic nerve (Supplementary Fig. 2B). Genome sequencing revealed a heterozygous c.965_980del p.(Ser322Ter), present in both the proband and unaffected parent. Proband (B1), presented with a severe form of bilateral microphthalmia with a vestigial remnant of eye, delayed motor development, intellectual disability, abnormal behavior, and schwannoma. This proband carried a heterozygous c.902G>A p.(Trp301Ter) variant. These two nonsense variants are expected to cause loss of protein function either through nonsense-mediated decay or truncation of the putative nuclear receptor ligand binding domain (NR-LBD, Fig. 1H). Proband D (Supplementary Table 4) had a disorder of sex development carried variant c.288dup p.(Cys96TrpfsTer4), which was absent in either parent. One of the parents, was also affected with a disorder of sex development, suggesting that the *NR6A1* variant is likely not associated with the condition.

The UK100KGP MAC cohort, which consists of 215 probands, was queried for rare missense and in-frame insertion/deletion variants. Proband C1, presented with bilateral microcornea and coloboma affecting the iris, choroid/retina, and optic nerve. One sibling had a similar condition by report. Both parents, and the two other siblings of the proband had no history of coloboma by report. Genome sequencing revealed a heterozygous variant c.227_229del p.(Ser76del) present in the proband (Fig. 1H, Supplementary Table 4). This variant leads to an in-frame deletion of a serine within the Zn-finger motif. Within the three MAC patients we report, no candidate pathogenic variants were found in the known MAC genes present in the current Genomics England PanelApp (ocular coloboma v1.47, anophthalmia or microphthalmia v1.51, structural eye disease v3.79). Thus, these cases further support that rare variants in *NR6A1* can cause MAC with reduced penetrance.

### Molecular Modeling Suggests Missense Variants Disrupt Important Intramolecular Interactions

The NR6A1 amino acid sequence is well-conserved between human, mouse, and zebrafish; specifically, the residues Ser76, Arg92 and Arg436 are conserved across multiple species (Supplementary Fig. 3). To understand the effects, missense variants had on protein stability and function, we created an *in silico* model of a complex of NR6A1 with DNA (Supplementary Fig. 4A). The AlphaFold model of NR6A1 is shown by the composition of Zn-finger (residues 60-172) and NR_LBD (residues 246-480) domains shown in orange and green, respectively. The rest of the model shown in gray is predicted as an irregular structure by AlphaFold. The locations of variants R92W and R436C are labeled. In wild-type (WT) NR6A1, a positively charged arginine residue 92 interacts with negatively charged DNA (Supplementary Fig. 4B). The R92W variant replaces the R92 residue with hydrophobic tryptophan (W), interrupting the electrostatic interaction with DNA and possibly reducing the DNA binding of the Zn-finger motif. The R436C variant affects the putative nuclear receptor ligand binding domain NR_LBD. In NR6A1, hydrogen atom 1HH2 of arginine R436 closely interacts with the oxygen atom of glutamic acid E388 (Supplementary Fig. 4C). The Variant R436C breaks this bond creating a cysteine residue instead of an arginine. In this variant domain, residues C443, C391, and C422 are distanced at 8-12 Å from C436. In the native state of protein, the reduced state of cysteine residues is protected. However, any oxidative damage could cause the formation of non-native disulfide bonds affecting protein structure and ligand binding.

### Missense variants alter NR6A1 protein subcellular localization

To study the functional impact of the missense variants on protein localization in the cell, the R92W and R436C mutations were introduced in WT *NR6A1* cDNA fused to a GFP coding sequence. All experiments were performed in context to the *NR6A1* isoform NM_033334.4 and repeated at least three times. Transfection efficiencies were between 50-60% and not significantly different between WT and variant constructs as analyzed by flow-cytometry and Western blotting (Supplementary Fig. 5, 6). The WT-*NR6A1* when over-expressed in HEK293 cells was consistently observed to localize in the nucleus (Fig. 2A, B), consistent with a previous report. The R92W variant, although nuclear, was not uniform in its distribution, forming apparent protein aggregates. In contrast, the R436C variant localized exclusively in the cytoplasm (Fig. 2A). The above-described localization pattern of the WT and variant isoforms was consistent in all transfected cells and across multiple rounds of transfection. Taken together these results suggest that both missense variants likely interfere with NR6A1 function due to improper subcellular localization.

**Fig. 2:**
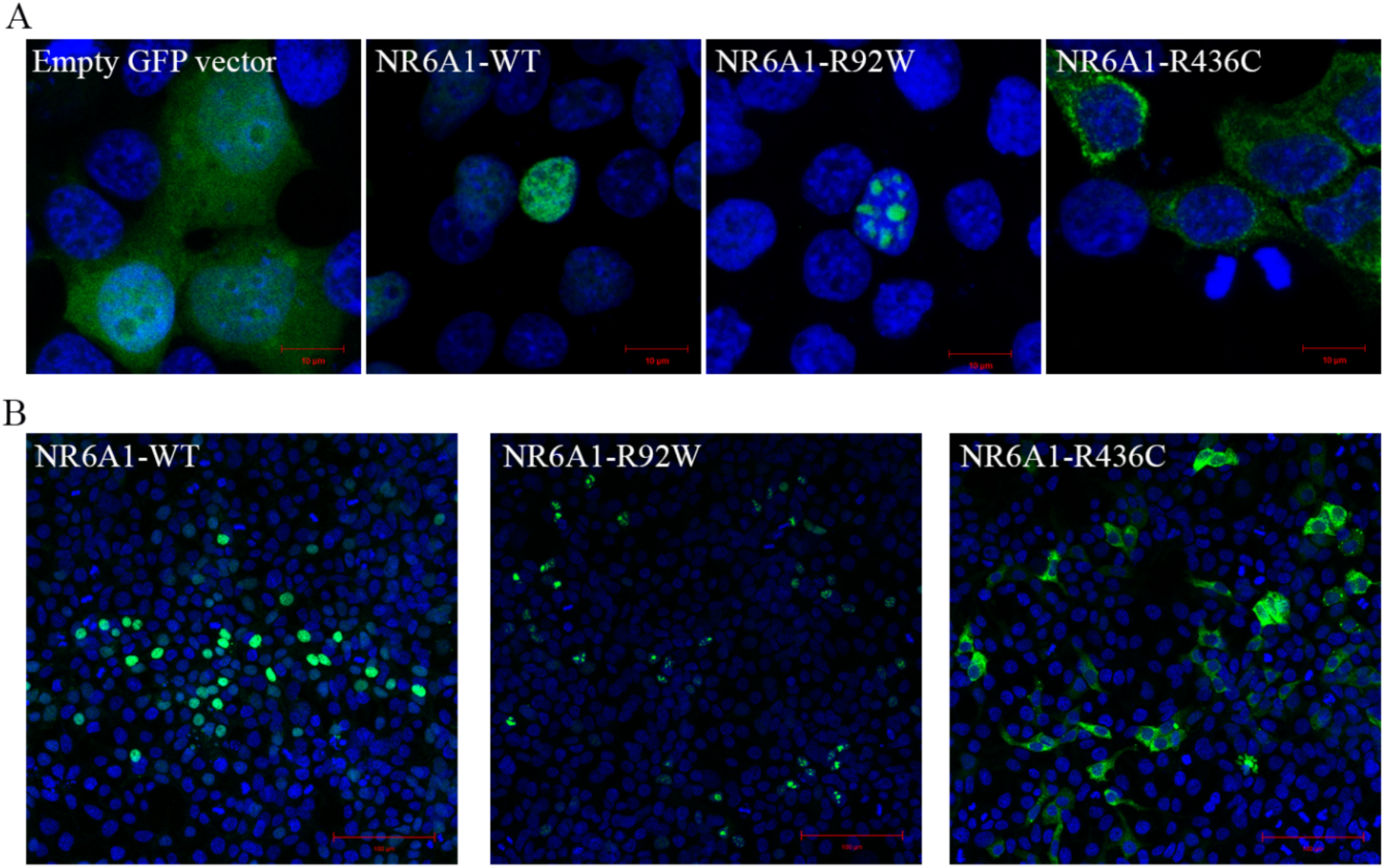
Subcellular localization of wild-type (WT) and mutant forms of NR6A1. NR6A1 variant localization pattern was studied by overexpression in HEK293 cells and representative high magnification (63X) images are shown from three different trials (A). Scale bar = 10 µm. The localization pattern for the WT and the two variant isoforms was observed to be consistent across three transfection experiments. (Cells counted: WT=387, R92W=350 and R436C=217). Scale bar = 100 µm.

### Expression pattern of mouse and zebrafish NR6A1 homologs suggests a role in early eye, kidney, and somite development

Analysis of bulk RNA-Seq datasets from ocular and non-ocular tissues demonstrates modest expression of *NR6A1* in most tissues and relatively higher levels of expression in embryonic stem cells/induced pluripotent stem cells (compared to adult ocular tissues) and in bone marrow and testis systemically (Fig. 3A, D)^23,24^. In the Human Retinal Cell Atlas single nucleus RNA-Seq dataset *, NR6A1* is highly expressed in adult horizontal cells and low in microglia and RPE (Fig. 3B)^25^. Expression of *NR6A1* is strongly correlated (>5 fold enrichment, p = 0.0024) with that of other coloboma-associated genes in fetal ocular tissues (Fig. 3C). This strength of enrichment was not seen in Genotype-Tissue Expression (GTEx) body tissue (p=0.361) or adult eye tissue (p=0.451) ^23,24^. We note that several of the enriched genes-*SALL4* (Duane-Radial Ray Syndrome), *PAX2* (Papillorenal syndrome), *ACGT1* (Baraitser-Winter Syndrome 2), *SALL1* (Townes-Brocks Syndrome 1) can also present with congenital renal anomalies.

**Fig. 3:**
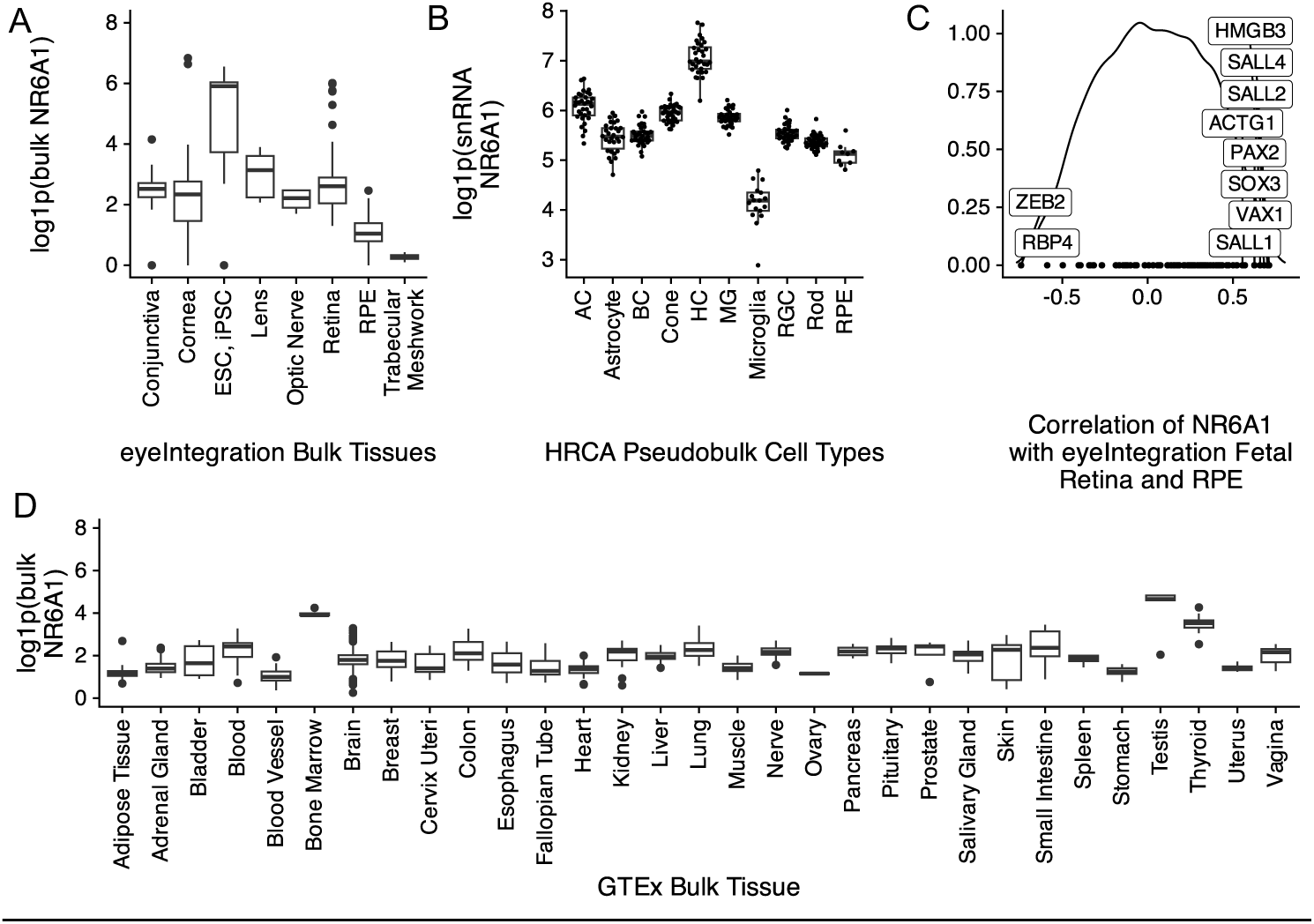
A. Comparative levels of NR6A1 from publicly available bulk human tissue RNA-sequencing (RNA-Seq) datasets accessed on the eyeIntegration website (https://eyeintegration.nei.nih.gov/). On average, expression is higher in embryonic and induced pluripotent stem cells (ESC, iPSC, respectively) than in adult ocular tissues. B. In adult retina, expression of *NR6A1* is highest in horizontal cells (HC) compared to other cell types in the Human Retinal Cell Atlas (HRCA) (AC, amacrine cell; BC, bipolar cell; MG, Müller glia; RGC, retinal ganglion cell; RPE, retinal pigment epithelium). C. Correlation of NR6A1 expression with fetal retina and RPE RNA-Seq data demonstrates association with several known coloboma associated genes (boxed labels). D. Among systemic tissues, *NR6A1* is expressed most highly in bone marrow and testes.

To establish plausible causation for *NR6A1* variants, we studied the embryonic expression of its orthologs in mouse and zebrafish model systems at developmentally relevant time points. Previous work has demonstrated widespread expression of *Nr6a1* in mouse at E8.5 and E9.5 (including the optic vesicle) that becomes nearly undetectable by E12.5. To study expression in the optic cup around the time of optic fissure closure, we used a probe that detects all validated transcripts of mouse *Nr6a1* at embryonic day 10.5 (E10.5, early optic cup) and E11.5 (time of optic fissure closure). At E10.5, we noted diffuse low-level expression throughout the early optic cup and surrounding tissues that becomes significantly decreased by the time optic fissure closure commences (E11.5) (Supplementary Fig. 7).

In zebrafish, *nr6a1* has two paralogs, *nr6a1a* and *nr6a1b,* both of which are maternally expressed. At 11hpf, when the optic vesicle evaginates, *nr6a1a* is widely expressed throughout the embryo, especially rostrally, showing less expression towards the posterior embryo axis (Fig. 4A). At 16hpf, *nr6a1a* remains widely expressed becoming restricted to the ventral regions of the brain, epiphysis, periocular tissues, heart and in the notochord and neural tube (Fig. 4B). Notably, *nr6a1a* expression is absent from the neural-mesodermal progenitor region in the tail of zebrafish embryos, consistent with its role in the trunk differentiation program. By 19hpf the expression appears to decrease overall but remains present in the ventral brain regions, notochord, somites, and the pronephric duct (Fig. 4C). At 24hpf, expression is prominent in the anterior diencephalon, tegmentum, midbrain, and along most of the length of the embryo in the neural tube; interestingly, expression is nearly absent from the neural retina and retina pigmented epithelia but is prominent in the lens (Fig. 4D-D’’), a pattern not noted in the mouse *Nr6a1* expression. After 26hpf and up to 72hpf we observed no detectable *nr6a1a* expression, consistent with published single-cell mRNA expression during zebrafish development.

**Fig. 4:**
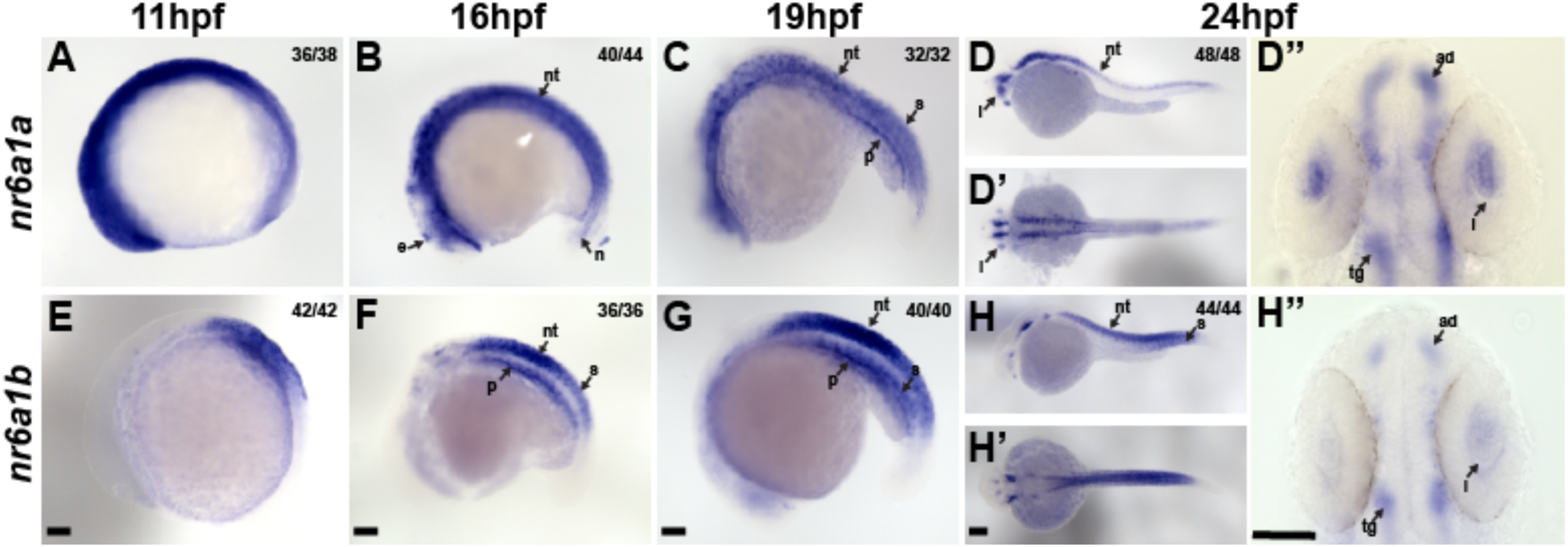
Expression pattern of *nr6a1a* and *nr6a1b* paralogs in zebrafish. *nr6a1a* is expressed ubiquitously at 11 hours post-fertilization (hpf) (A). By 16-19 hpf (B, C) expression is present in the somites (S), neural tube (NT), and notochord (N). At 24 hpf, expression remains in the NT but is decreased in the S and N. Expression in the lens (L) is first noted at 19 hpf and is particularly prominent by 24 hpf (D-D’’). *nr6a1b* expression at 11 hpf is anterior trunk, localizing to neural tube and somites from 16 hpf (F) and 19 hpf (G). At 24hpf (H-H’’) it remains expressed in the neural tube and somites, with faint expression can be seen in the lens. All embryos are oriented in a lateral view, anterior to the left and dorsal up, except D’ and H’ shown in dorsal views. Scale bar = 100 µM. e-epiphysis, l-lens, p-pronephros, n-notochord, s-somite, nt-neural tube, ad-anterior diencephalon, tg-tegementum.

Unlike *nr6a1a*, *nr6a1b* expression at 11hpf is limited to a patch in the posterior neuroectoderm of the embryo but excluded from the most caudal region (Fig. 4E). At 16hpf and 19hpf, *nr6a1b* expression is prominent in the neural tube, somites, and pronephric duct and, like *nr6a1a,* is excluded from the neural-mesodermal progenitor region in the tail (Fig. 4F, G). By 24hpf, expression is decreased in most tissues but remains in the tegmentum, cranial ganglia, neural tube, and somites in the distal region of the trunk (Fig. 4H-H’’). By 36hpf and through 72hpf, *nr6a1b* is notably expressed in the developing lens, brain, and cranial ganglions. (Supplementary Fig. 8).

### Morpholino knockdown of zebrafish nr6a1a/nr6a1b recapitulates human phenotypes which are not rescued by pathogenic variant mRNA

All the morpholinos experiments are carried out following the guidelines set forth for their use in zebrafish. To test the functional consequences of *nr6a1a* and *nr6a1b* knockdown, we designed translation (TB) and splice blocking (SB) morpholinos for each paralog of the gene. Morphants were divided into four phenotypes: normal, mild (normal/near normal body axis w/ microphthalmia), moderate (slightly shortened and mildly curved body axis, microphthalmia ± coloboma and heart edema) or severe (significantly shortened and curved body axis, microphthalmia ± coloboma, heart edema) (Fig. 5A-D, Supplementary Fig. 9, 10). Embryos were scored at 72 hpf (after optic fissure closure and initial stages of eye growth are normally completed) to ensure microphthalmia/coloboma represents a true phenotype and not because of developmental delay or undergoing growth compensation.

**Fig. 5:**
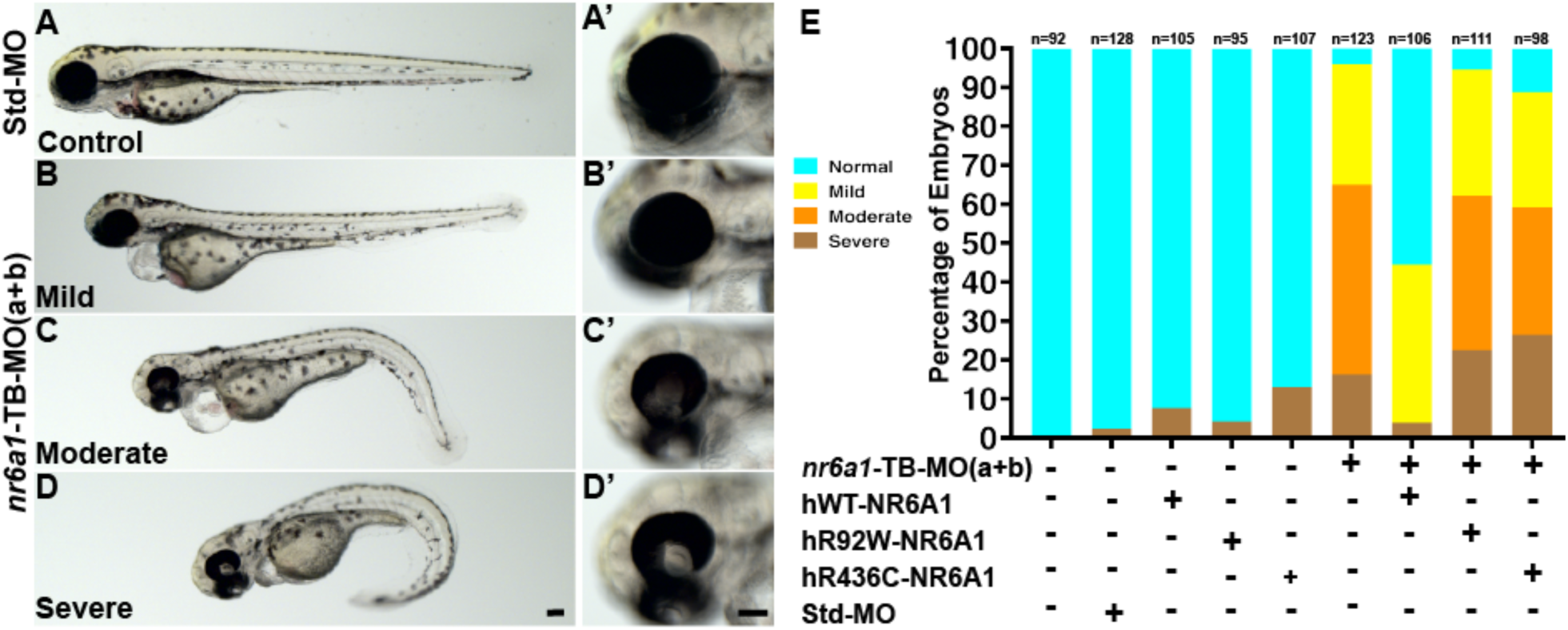
Rescue of *nr6a1+nr6a1b* zebrafish morphant phenotypes with wildtype and mutant human NR6A1 mRNA: Controls (A, A’) have a straight body axis and the optic fissure (OF) is closed. The ***nr6a1+nr6a1b*** morphants that have a mild phenotype (B, B’) have close to a normal body with microphthalmia and heart edema; a moderate phenotype (C, C’) with a slightly bent body axis with smaller eyes, coloboma and a severe heart edema; and severe morphants (D, D”) have a curved body axis with smaller eyes, coloboma and heart edema. The morphant phenotype was rescued when the morpholinos were co-injected along with the human-*NR6A1*-wild type mRNA. However, there was no significant rescue in the morphant phenotype when the morpholinos were injected with either R92W or R436C human disease-causing variants (E). Morpholinos were injected at 0.75 ng each (1.5 ng total). Scale bar = 100µM

Knockdown of *nr6a1a* (Supplementary Fig. 9E) or *nr6a1b* (Supplementary Fig. 10E) with either TB-MO or SB-MO resulted in a significant number of moderate/severe phenotypes with few mild phenotypes. Although the effect of TB-MO and SB-MO were similarly potent for *nr6a1b* knockdown, the SB-MO had a stronger effect than the TB-MO for *nr6a1a*. SB-MO knockdown of the gene was validated for both paralogs by reverse transcription-PCR experiments (Supplementary Fig. 9F, 10F). The phenotypic spectrum was not affected by co-injection with p53 morpholino, suggesting widespread cell death was not the primary cause of our observations (data not shown).

Overexpression of 100 pg of human *NR6A1* mRNA in zebrafish shows no overt phenotype (Supplementary Fig. 11A, B). Co-injection of 2 ng and 1.25ng of nr6a1a and nr6a1b, TB-MO respectively along with 100 pg of WT human mRNA (hWT-*NR6A1*), resulted in a rescue, with over 60% embryos exhibiting a normal/control-injected phenotype (Supplementary Fig. 9G, S10G). In contrast, co-injection with either hR92W or the hR436C missense variants of *NR6A1* identified in coloboma patients were significantly less effective in rescuing the zebrafish *nr6a1a/b* knockdown, indicating that the missense variants are deleterious (Supplementary Fig. 9G, 10G).

To study the effect of knocking down both *nr6a1a* and *nr6a1b* zebrafish paralogues, we co-injected 0.75 ng of TB-MO for each paralog (1.5 ng total), resulting in a similar spectrum of phenotypes compared to the knockdown of individual paralogues (Fig. 5A-D, A’-D’). While injection of TB-MO resulted in >60% embryos having a moderate or severe phenotype, co-injection of 100 pg hWT-*NR6A1* mRNA, resulted in >50% normal embryos. Neither the hR92W or hR436C *NR6A1* mRNAs resulted in significant rescue, confirming the pathogenicity of these variants (Fig. 5E). Injection of 0.75 ng of either *nr6a1a* TB-MO or *nr6a1b* TB-MO resulted in a significantly milder phenotype, suggesting that co-injection of these had at least an additive phenotypic effect in the combined MO injection experiment (Supplementary Fig. 12).

Because a prior study reported that both overexpression and loss-of-function of *nr6a1* can result in developmental phenotypes in *Xenopus laevis*, we also evaluated the effect of injection of human *NR6A1* mRNA on zebrafish development. Overexpression of 150 pg of *hNR6A1* mRNA resulted in microphthalmia and heart edema with a straight body axis (n=91/108) (Supplementary Fig. 11C). At 200 pg, overexpression of *hNR6A1* mRNA, phenotypes were more severe including colobomatous microphthalmia, heart edema and a bent body axis (n=60/92), with 26% (n=24/92) exhibiting noticeable shortening and loss of chevron-shaped somites (Supplementary Fig. 13); a minority of embryos (n=8/92) developed no discernible eyes (Supplementary Fig. 11F). Taken together, these experiments demonstrate that normal zebrafish eye development is sensitive to *nr6a1* dosage and both reduced and increased *nr6a1* expression result in developmental phenotypes analogous to human colobomatous microphthalmia.

## Discussion

Here we describe six *NR6A1* variants that cause an autosomal dominant syndromic form of colobomatous microphthalmia and missing vertebrae with or without congenital kidney abnormalities, that we term OVR syndrome. As with many other cases of syndromic and non-syndromic microphthalmia/coloboma, the OVR syndrome show incomplete penetrance and variable expressivity ^1^. By 2015 ACMG/AMP variant interpretation criteria, we considered chr9:g.124536516_124643457del pathogenic (criteria: PVS1, PP1_Strong, PM2) and other MAC-associated variants likely pathogenic (criteria: Ser76del, PM1, PM2, PM4, PP3; Arg92Trp, PS3, PM1, PM2, PP3; Arg436Cys, PS3, PM2, PP3; Ser301Ter & Ser322Ter, PVS1, PM2). Thus, *NR6A1* variants were causative among 1.3% - 1.4% families in two independent patient cohorts (3 out of 224 in the NEI coloboma cohort and 3 out of 215 in the MAC cohort in the UK100KGP).

The NEI study, which specifically recruits patients with coloboma/microphthalmia, performs extensive phenotypic analysis on probands including complete eye examination, kidney ultrasound, neuropsychological testing, physical exam/dysmorphology exam, spine x-ray, routine bloodwork/urinalysis, ECHO (in the presence of a murmur), and audiology. Additional testing (e.g., brain MRI) may be performed on an as needed basis. In addition, all available first-degree relatives undergo a complete dilated fundus exam. As such, we have greater certainty that a patient is truly unaffected, say, by coloboma, rather than being simply asymptomatic. Indeed, the mother of the proband in family COL034 (COL034.2), for example, was visually asymptomatic and unaware of a *forme fruste* of coloboma or a missing thoracic vertebra prior to her exam with us. Conversely, the Genomics England database spans an entire population in a gene and phenotype agnostic manner but may contain incomplete or unrelated phenotypic information. As such, phenotypes such as intellectual disability (Individual B1, Table S4) may be spurious associations or may be uncommon manifestations of an *NR6A1*-related syndrome. Confirmation of these and other possible phenotypes awaits description of additional cases. We include congenital renal disease as part of this new syndrome not only because two individuals in two separate pedigrees exhibited these phenotypes, but also because Rasouly et al. have simultaneously identified presumed loss-of-function variants in thirteen individuals with congenital renal abnormalities, with or without congenital eye abnormalities, providing further validation of our findings (personal communication).

The genotypes and functional data we present suggest haploinsufficiency as the primary mechanism of disease, although we cannot rule out that missense variants may have other, dominant-negative effects, by dimerizing with the wildtype protein or interacting with other transcription regulators. The differences in subcellular localization of the two missense variants in *NR6A1* may indeed hint at more than one mechanism of disease.

The early expression of *NR6A1* homologs in mouse and zebrafish are consistent with the previous data and suggest that the colobomatous microphthalmia observed in our patients may result from effects on early eye morphogenesis rather than a defect in optic fissure closure *per se*. However, given the expression of *nr6a1a/nr6a1b* in the lens vesicle in zebrafish, a non-cell autonomous effect on optic fissure closure cannot be excluded. In fact, evidence from Mexican surface and cave fish (*Astyanax mexicanus)* experiments show that early neural retina development and maintenance relies on a healthy lens ^26,27^.

Recently, *NR6A1* has been shown to be important for somite development and, consequently, vertebral number, thus strengthening the phenotyping link with missing vertebrae we describe in humans^19–22,28^. Vertebrae differentiate from somites which develop their stereotyped segmentation pattern in an anterior to posterior progression during early development, with successive *HOX* genes specifying different regions of the spine via a process called temporal collinearity. Homozygous germline inactivation of *Nr6a1* in mice results in embryonic lethality around E10.5 with cardiovascular, neural tube and hindgut abnormalities as well as fewer somites (13, rather than the normal 25). In S*us domesticus* (pig), *NR6A1* was identified as a quantitative trait locus for vertebral number, which is known to vary between breeds^19,21^. In *Equus assinus* (donkey), an *NR6A1* intronic polymorphism is associated with body size/vertebral number and a single nucleotide polymorphism in exon 8 is associated with the number of lumbar vertebrae in Kazakh sheep ^20,28^. In developing *Xenopus*, *NR6A1* is expressed in late tailbud and neurula stages; overexpression results in posterior defects and disturbed somite formation, while expression of a dominant negative form of the receptor results in abnormal neural tube differentiation, loss of head structure including eyes, and downregulation of a retinoic acid receptor (RARγ2) anteriorly. Retinoic acid treatment of embryos upregulates expression of NR6A1, increasing primary neurogenesis via factors such as NeuroD, XDelta1 and x-ngnrl. Retinoic acid is a known and important regulator of both ocular and kidney development ^29,30^; whether retinoic acid receptor signaling is disrupted in model systems of *Nr6a1/nr6a1* is currently under investigation. However, all phenotypes previously observed when modulating the activity of *NR6A1* in animal models are consistent with the developmental defects in the eyes, kidneys, and vertebrae that we observe in patients carrying deleterious mutations in NR6A1.

In conclusion, genome sequencing identified novel *NR6A1* variants in three unrelated families which are associated with a novel OVR syndrome, these findings were further corroborated in an independent cohort using a genome first approach. Using *in silico* prediction and molecular studies we demonstrated that these highly conserved variants disrupt NR6A1 protein structure leading to mis-localization at the cellular level. We further demonstrated enrichment of coloboma-associated genes with *NR6A1* in fetal, but not adult tissues. Expression of *NR6A1* homologs in mouse and zebrafish embryos suggests disease relevant tissue-specific gene expression pattern. This was further confirmed by *in vivo* experiments where the knockdown of zebrafish *nr6a1a and nr6a1b* resulted in ocular and systemic phenotypes that were partially rescued with WT human *NR6A1* mRNA but not with the two variants tested. This data implicates the human *NR6A1* gene variants with the OVR syndrome.

## Data Availability

All data produced in the present work are contained in the manuscript and UK100KGP data was managed by Genomics Data England Limited.

## Acknowledgment

We are grateful to the patients and their families for participation in this research. We thank the staff at the NIH Clinical Center and the NEI Eye Clinic for patient phenotyping, ophthalmic testing, and ophthalmic imaging. We are grateful to Drs. Hila Milo Rasouly, Krishna Murthy, Ali Gharavi for their critical reading of this manuscript and for facilitating publishing our similar findings in parallel. We thank Dr. Amy Hodson-Thompson for her expert assistance in designing and formatting figures, and Dr. Thomas Hawkins and Dr. Gareth Powell for advice on the interpretation of zebrafish expression patterns. We thank Dr. Robert Fariss and members of the NEI Imaging Core for use of their confocal imaging facility and Dr. Elena Semina for helpful discussions on this project.

This research was made possible through access to data in the National Genomic Research Library, which is managed by Genomics England Limited (a company owned by the Department of Health and Social Care). The National Genomic Research Library holds data provided by patients and collected by the NHS as part of their care and data collected as part of their participation in research. The National Genomic Research Library is funded by the National Institute for Health Research and NHS England. The Wellcome Trust Cancer Research UK, and the Medical Research Council have also funded research infrastructure. This research was supported by the NIHR Moorfields Biomedical Research Centre (GA, MM, SL).

## Authors Contribution

EU, RBH, MIM, GA, RY and BG performed exome/genome analysis and variant confirmation in the NIH and UK100KGP populations. RA, AN and CB performed variant confirmation, including confirmation of deletion breakpoints. IHM, DB, SL, TGT, MM and BPB performed patient examination/phenotyping. RY and MIM provided phenotypic data from the UK100KGP. YVS performed molecular dynamic modeling. DM performed bioinformatic analyses of *NR6A1*. UMN and DSM performed zebrafish *in situ* hybridization, morpholino knockdown and mRNA rescue experiments. RY assisted in interpretation of zebrafish data. AG and CA performed cell culture and transfection experiments. AG performed confocal microscopy. RV performed flow cytometry. All co-authors provided draft language and experimental design for their portions of the manuscript, as well as a critical review of the entire manuscript. BPB provided overall project conception and experimental design, drafting of the manuscript and project support.

## Funding

This study was supported by the Intramural Research Program of the NIH. GA is supported by a Fight For Sight (UK) Early Career Investigator Award (5045/46), the National Institute of Health Research Biomedical Research Centre (NIHR-BRC) at Moorfields Eye Hospital, and the UCL Institute of Ophthalmology, and Moorfields Eye Charity (Stephen and Elizabeth Archer in memory of Marion Woods) and NIH-P20GM139769. RY was supported by the Moorfields Eye Charity Career Development Award and Springboard (GR001155 and GR001210), Medical Research Council (MR/X001067/1) and FODNECYT (1221843). MIM was supported by Moorfields Eye Charity PhD Studentship (GR001661).

---

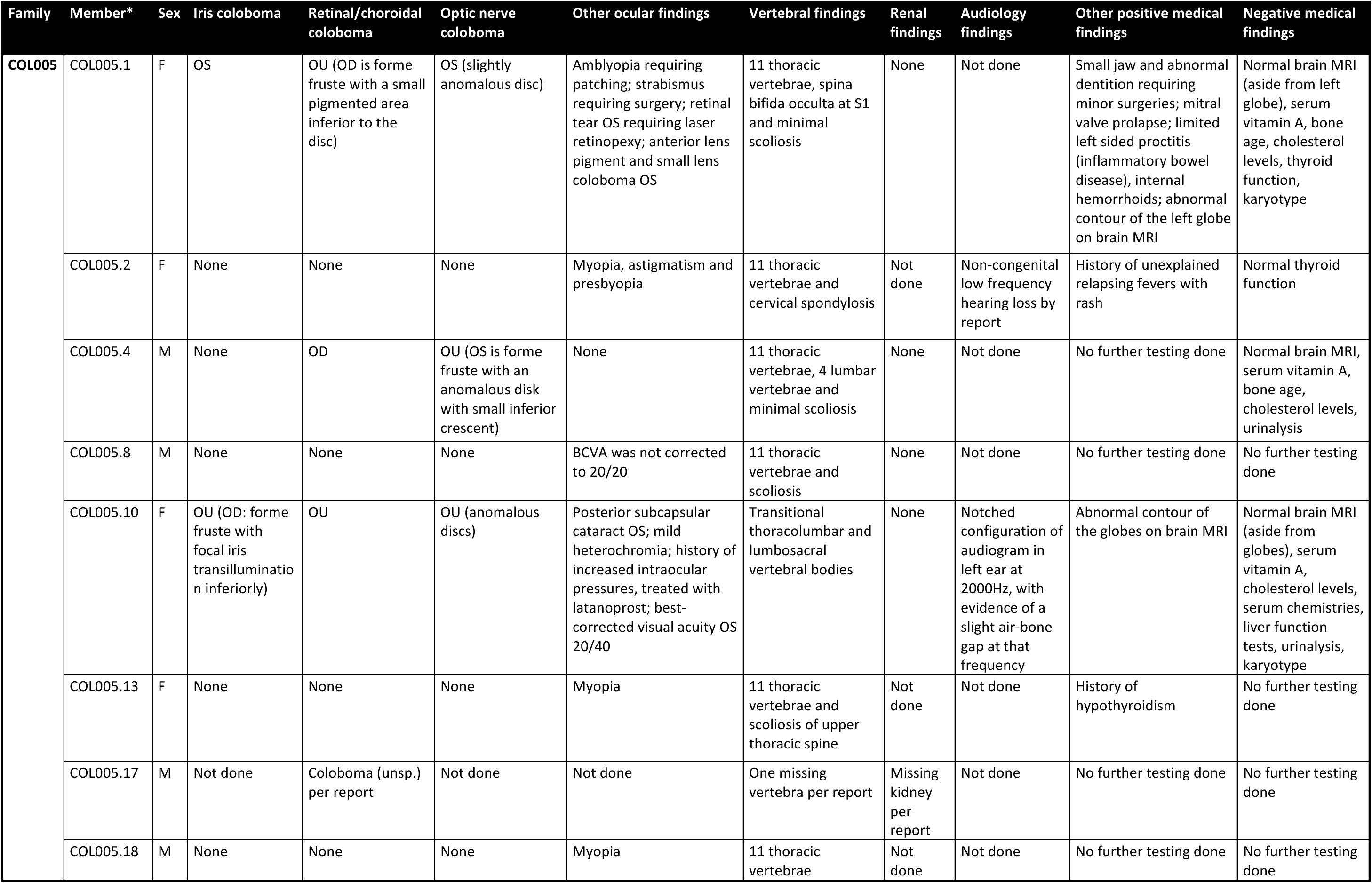
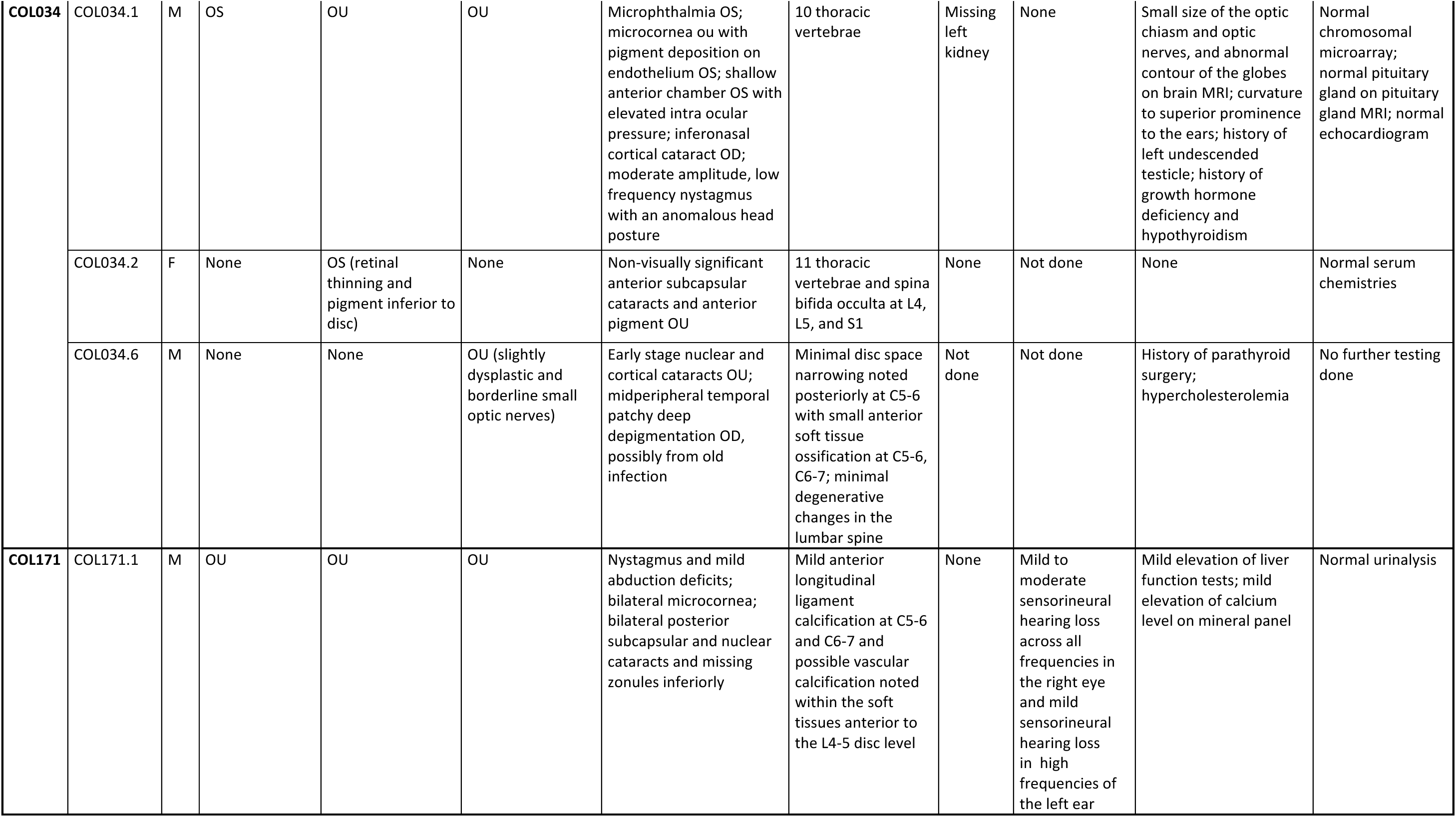

## Extended Data

### COL005

The proband of family COL005 (COL005.1) presented with bilateral uveal colobomas (Figure 1B, C). Past ocular history was remarkable for strabismus surgery and amblyopia OS treated with patching. Past medical history was notable for a “small jaw requiring minor reconstruction for proper dentition”, “abnormal enamel to teeth, requiring capping”, and speech therapy. Family history was notable for a sibling (COL005.4) and relatives (COL005.10), (COL005.17) with uveal coloboma. Best-corrected visual acuity was 20/15 OD and 20/60-OS (Snellen). Ocular motility was full with small exophorias at distance and near. Pupils were remarkable for a left iris coloboma. Dilated fundus examination showed a linear pigment disturbance inferior to the optic nerve OD (likely a *forme fruste* of coloboma) and a large chorioretinal coloboma OS inferior to the nerve and macula with hyperpigmentation in its periphery. Several years later, a retinal tear developed in the inferior periphery OS that was treated with laser and has remained stable. Systemic examination was remarkable for 11 thoracic vertebrae, mild scoliosis and spina bifida occulta of S1 on spine x-ray. Mitral valve prolapse was noted on ECHO. Kidney ultrasound, brain MRI, bone age, serum cholesterol, vitamin A, karyotype/sub-telomeric FISH and thyroid function were normal. Genome sequencing in two distantly related family members COL005.1 and COL005.10 revealed the same heterozygous 107kb deletion in *NR6A1* (chr9:g.124536516_124643457del, GRCh38). The deletion breakpoints were in intron 2 and 6 removing the coding sequence for amino acids (aa) Ile48-Gly275 and likely causing a frameshift (p.Ile48Asnfs*3, Figure 1H). The status of the heterozygous deletion was determined by breakpoint PCR among family members available, which revealed complete segregation with the missing vertebra with an estimated LOD score of 3.6 (Figure 1A, Figure S1). Four family members were also affected by coloboma in addition to missing vertebra, of which one (COL005.17) also had, by report, only one kidney.

### COL034

The proband of family COL034 (COL034.1) presented with bilateral uveal colobomas, microphthalmia OS (Figure 1E) and a moderate amplitude, low frequency nystagmus with an anomalous head posture. Prenatal history was remarkable for maternal oral contraceptive use at time of pregnancy and a prenatal ultrasound that showed a two-vessel umbilical cord and inability to visualize the left kidney. Delivery was at term and unremarkable with a birth weight of 5lbs, 3oz. Subsequent growth was slow, requiring growth hormone injections. Ocular examination was remarkable for bilateral microcornea (horizontal diameter 9mm OD, 6mm OS), a left iris coloboma, left sensory esotropia, an inferonasal cortical cataract OD, and bilateral chorioretinal and optic nerve colobomas. Best-corrected visual acuity as a kid was 20/400 OD and <20/800 OS by ETDRS chart. By teens, the left lens began to dislocate slightly and develop mild/moderate nuclear opacity. Spine x-ray demonstrated 10 thoracic vertebrae. Kidney ultrasound showed a normal right kidney and a missing left kidney (normal retroperitoneal or pelvic). Physical exam was remarkable for curvature to superior prominence to the ears, and a left undescended testicle. Chromosomal microarray was normal. Audiology examination and an echocardiogram were unremarkable. Subsequent endocrine workup showed hypothyroidism requiring replacement (in addition to his growth hormone deficiency). In late teens, developed intraocular pressures in the low/mid-twenties with a history of slowly progressive vision loss to hand motion, prompting initiation of intraocular pressure lowering drops. A brain MRI showed small optic nerves/chiasm, no parenchymal abnormalities, and a normal-appearing pituitary. Genome sequencing revealed a heterozygous c. 274C>T p. (Arg92Trp) variant in *NR6A1*, which was found in the affected mother and essentially unaffected grandfather (Figure 1A, H).

### COL171

The proband of family COL171 (COL171.1) presented with bilateral colobomatous microphthalmia affecting the iris, retina/choroid and optic nerve. Visual acuity was 20/640 OD and 20/250 OS. Nystagmus and mild abduction deficits were noted. Slit lamp exam was notable for bilateral microcornea, bilateral posterior subcapsular and nuclear cataracts and missing zonules inferiorly OU (Figure 1F). By report, kidney ultrasound, ECHO and physical exam were normal at birth. Spine x-ray showed normal number and morphology of vertebrae. Audiology examination was notable for mild to moderate sensorineural hearing loss across all frequencies in the right ear and mild sensorineural hearing loss in high frequencies of the left ear. Blood chemistries notable for mild elevation of liver function tests and calcium. Genome sequencing revealed a heterozygous c.1306C>T p.(Arg436Cys) variant in the proband which was absent in his unaffected mother (Figure 1H). Additional family members were not available for segregation analysis, but his father had no history of coloboma by report.

## Supplementary Methods

### Genetic testing

Genomic DNA samples prepared from blood or saliva from NEI patients, and their family members were subjected to short-read Next-generation sequencing (NGS) using Illumina platforms. In total, 101 proband samples were subjected to amplicon sequencing of the *NR6A1* gene (Table S1) using a MiSeq sequencer (2 x 300 bp paired-end), 57 samples subjected to exome sequencing (2 x 150 bp paired-end, xGen exome v1 supplemented with additional probes, Blueprint Genetics), 66 samples subjected to GS (2 x 150 bp paired-end, PCR-free library, NIH Intramural Sequencing Center). Reads were aligned to the GRCh38 reference genome, small variants and structural variants were then called, annotated, and prioritized using a custom NGS analysis pipeline (https://github.com/NIH-NEI/NGS_genotype_calling & https://github.com/NIH-NEI/variant_prioritization).

Sanger sequencing was performed to confirm select variants in probands and family members using the BigDye-direct sequencing kit (Thermo Fisher) using primers provided in Table S1. The deletion breakpoint in family COL005 was also determined by PCR and Sanger sequencing (Table S1). Breakpoint PCR was further used for genotyping of the COL005 family. The logarithm of the odds (LOD) score in family COL005 was estimated using the formula log_10_(1/0.5^Segregations^).

Additional patients and family members underwent Genome Sequencing (GS) as part of the UK100KGP including the clinical variant interpretation pipeline (The National Genomics Research Library v5.1, Genomics England. doi:10.6084/m9.figshare.4530893/7. 2020.). Genome data from affected individuals recruited with a clinical phenotype in keeping with microphthalmia, anophthalmia or coloboma were interrogated for rare (minor allele frequency <0.001, gnomAD v3.1 dataset) biallelic or *de novo* protein altering variants across the genome. Candidate variants underwent manual curation including *in silico* prediction, literature search and pathway analysis to establish biological plausibility as a pathogenic variant in developmental eye disease. Additional analyses of all rare protein altering variants in *NR6A1* across the entire UK100KGP was performed to identify any individuals outside of the ophthalmology cohort who harbored a candidate pathogenic variant. All variants were manually inspected in the Integrative Genomics Viewer (IGV) after loading sample bam files. Variants appeared to be artifacts were not reported.

### Variant classification

The 2015 ACMG/AMP sequence variant interpretation guidelines were followed for variant classification ^31,32^. The PM1 (functional domain) criterion was applied to variants in part of the DNA binding domain, a.a. Thr68-Lys119, as the region is highly constrained for missense variations in gnomAD (v2.1.1, missense observed/expected = 0.19, p-value = 6x10^-6^). The PP3 criterion was applied to missense variants based on a collection of in-house in silico prediction tools (https://github.com/NIH-NEI/variant_prioritization) and the inframe deletion variant based on five in silico prediction tools (CAPICE, FATHMM-indel, MutationTaster, MutPred-Indel, and SIFT.

### Molecular modeling

A structural model of NR6A1 was generated using the AlphaFold server, AF-Q15405-F1-model_v4). The Zn-finger domain (ZFD) and nuclear receptor ligand binding domain (NR_LBD) were saved as two PDB files. The binding of DNA to the ZFD of NR6A1 was modeled using a single ZFD domain of the retinoid X receptor alpha-liver X receptor beta (PDB ID: 4NQA) in a complex with DNA. Two variants (R92W and R436C) were generated using the Edit > Swap > Residue function on the respective domain PDB files in YASARA (http://www.yasara.org/). Variant models were optimized and minimized using gradient descent. All two minimized mutants and the two WT, ZFD and NR_LBD models were subjected to 10 ns of Molecular Dynamics (MD) using YASARA’s ‘run.mcr’ macro. Ion concentration was added as a mass fraction with 0.9% NaCl. The simulation temperature was set to 310 K with a water density of 0.997 g/mL. For each domain, the cell size extended to 10 Å beyond each side of the protein in the shape of a cube. Dimensions were 90.2Å x 90.2Å x 90.2 Å and 82.5 Å x 82.6 Å x 82.6 Å for the nuclear receptor Zn-finger and ligand-binding domains, respectively. Each simulation was run in YASARA using an AMBER14 forcefield, with a timestep of 2.5 fs. Simulation snapshots were outputted for every 0.1 ns, resulting in 100 simfiles for each simulation.

### Fish maintenance and zebrafish strains

*Danio rerio* were maintained under standard conditions. Embryos were staged according to Kimmel et al., 1995 ^33^. ABTL stocks were used for all the experiments, which were carried out in accordance with National Eye Institute, Animal Care and Use Committee Protocol Number NEI-648.

### Zebrafish *in situ* hybridization

Embryo were fixed in 4% paraformaldehyde (PFA) overnight at 4°C and dehydrated in methanol for 1h at −30°C. The embryos were rehydrated, treated with proteinase-K and re-fixed with 4% PFA. Pre-hybridization and hybridization were carried out at 65°C. RNA probes were synthesized using a DIG labeling kit (Millipore-Sigma, 112770739) following manufacturer’s protocol. *nr6a1a* RNA probe was synthesized from a CDS clone in TOPO TA vector (ThermoFischer Scientific), while *nr6a1b* was synthesized using PCR product as a template. Primers are noted in Table S2. Samples were hybridized overnight with RNA probes at 65°C, washed, incubated with Anti-DIG antibody (Millipore-Sigma, 1109327490); color was developed using BCIP/NBT substrate (Millipore-Sigma, 11681451001) in alkaline phosphatase buffer. Embryos were imaged with Leica DM6 dissecting microscope.

### Morpholino gene knockdown and rescue experiments in zebrafish

All morpholinos (MO) were obtained from Gene Tools LLC. MOs used to target zebrafish *nr6a1a* and *nr6a1b* are given in Table S3. Human *NR6A1*-wild type, variants *NR6A1*-R92W and *NR6A1*-R436C DNA fragments were synthesized and cloned in pCS2+ (Azenta Life Sciences). Plasmids were linearized with *Not I* restriction enzyme and capped mRNA was synthesized using mMessage mMachine T7 Transcription kit (ThermoFischer Scientific). MOs and mRNA were co-injected into zebrafish embryos at single cell stage. *nr6a1a* and *nr6a1b* translation blocking (TB) MOs were used at 2ng and 1.25ng respectively. Nr6a1a and nr6a1b, SB-MOs were injected at 2ng and 1ng respectively. Human *NR6A1*-wild type was used at 100pg and 150-200pg for RNA rescue and over expression studies respectively. *NR6A1*-R92W and *NR6A1*-R436C RNAs were used at 100pg for rescue experiments. For over-expression experiments, doses of 100pg-200pg *hNR6A1* mRNA were injected at the single cell stage. Embryo phenotypes were scored and imaged at 72 hours post-fertilization (hpf) using Leica DM6 dissecting microscope.

### Cell culture and transfection studies

HEK293T cells maintained in DMEM with 10% FBS and 1% penicillin-streptomycin were seeded onto 4-well chamber slides, maintained for 24 hr and transiently transfected with GFP tagged WT and/or mutant *NR6A1* constructs (Azenta Life Science, Burlington, MA, USA) using X-treme Gene HP (Roche, Indianapolis, IN, USA) following manufacturers’ instructions. After 24-48 hrs of transfection, transfected cells were fixed for 15 mins in 4% paraformaldehyde (PFA) in PBS. After washing with 1× PBS cells were incubated for 1 hr at room temperature with Hoechst33342 (1:250 dilution in PBST). Subsequently, the slides were washed and mounted with Fluoromount-G® (SouthernBiotech, Birmingham, AL, USA). Zeiss confocal microscopes 880 coupled with an Airyscan® detector was used for confocal imaging. The images were analyzed using ZEN Software (Carl Zeiss Microscopy LLC, Thornwood, NY). The cell culture experiments were repeated at least three times for each for variant localization studies.

### Flow cytometry

Transfection efficiency was determined by measuring the expression of GFP after 48 hrs post transfection. HEK293 cells were detached from the plates using Trypsin for 5 mins followed by neutralization with serum containing media. The cells were then fixed for 15 mins in 4% paraformaldehyde (PFA) in PBS and then collected in 1xPBS containing 2% FBS (FACS buffer) and washed 2 times by centrifugation. The cell suspension was filtered through a 50 µm cell strainer. Data was acquired with a CytoFlex NUV instrument (Beckman Coulter, Brea CA) using the blue light excitation and 525 nm emission to detect GFP and violet light excitation and 450 nm emission to detect DAPI detection. Data analysis was done using CytExpert software Version 2.5 (Beckman Coulter, Brea CA). Interesting cells were identified as DAPI negative, in the whole cell cluster in a FSC vs. SSC plot and being in a single cell state in the FSC-A vs. FSC-Width. Transfection efficiency was quantified as the Stain Index of GFP fluorescence intensity, which was calculated using the median fluorescent intensity and robust Standard Deviation as described.14 The cell culture experiments were repeated at least three times for each for variant localization studies.

### Mouse Embryo *in situ* Hybridization

*Nr6a1* mRNA expression in mouse was assayed by RNA *in situ* hybridization with *Nr6a1*(Cat: 1314941-C1) probe using the RNAScope Assay, Multiplex fluorescent Reagent Kit V2 (Advanced Cell Diagnostics (ACD), Newark, CA, USA) on E10.5 and E11.5 cryosection as previously described ^34^.

### Gene expression analysis of *NR6A1*

The h5ad (d27a79a1-8a5f-404d-8063-52e19122ef49.h5ad for adult and 88444d73-7f55-4a62-bcfe-e929878c6c78.h5ad for fetal) from the HRCA project were downloaded from cellxgene.cziscience.com and the raw counts were summed at the sample and cell type level to create a pseudobulk matrix with the python package ADPBulk (https://github.com/noamteyssier/adpbulk). The eyeIntegration (which includes GTEx) gene counts and metadata were downloaded from eyeIntegration.nei.nih.gov (https://hpc.nih.gov/~mcgaugheyd/eyeIntegration/2023/gene_counts.csv.gz and https://hpc.nih.gov/~mcgaugheyd/eyeIntegration/2023/eyeIntegration23_meta_2023_09_01.built.csv.gz).

The pseudobulk and bulk RNA-seq counts were normalized with by CPM and transformed in R/4.3 to have a mean of zero and a standard deviation of one. The first four principal components were removed with the WGCNA tool removePrincipalComponents. The correlation matrix was created with the base R cor function. The correlation scores were expression transformed with the spqn package’s normalize correlation function. Plots of the expression of NR6A1 were created in R/4.3 with the ggplot2, cowplot, and ggbeeswarm packages.

## Supplementary Tables

[See Excel Spreadsheet]

Supplementary **Table 1**: PCR primers used for human DNA sequencing

Supplementary **Table 2:** Primers used in zebrafish in situ experiments

Supplementary **Table 3:** Morpholinos used in zebrafish gene knock-down experiments.

Supplementary **Table 4:** Variant and phenotypic information of rare NR6A1 variant carriers in the UK100KGP cohort

Supplementary **Table 5:** Detailed breakdown of zebrafish morpholino experiments

## Supplementary Figures and Legends

**Supplementary Fig. 1:**
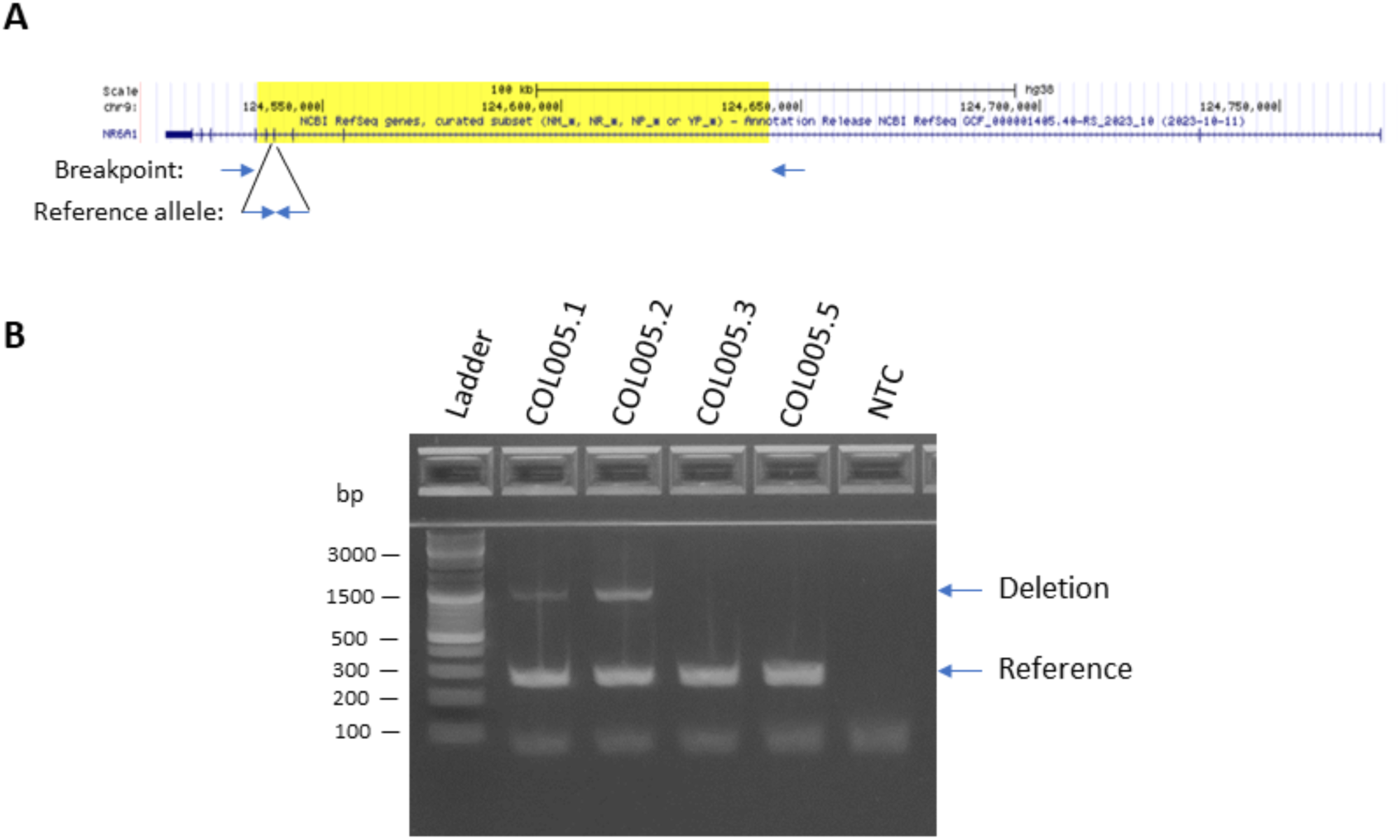
NR6A1 genotyping for the deletion in family COL005. (A), Diagram of the 107-kb deletion (yellow-highlighted) and primer binding sites. (B), Example image of the agarose gel electrophoresis of duplex PCR.

**Supplementary Fig. 2:** Phenotypes associated with *NR6A1* variants in UK100KGP. **A**. Pedigrees of three families (A; B; C) from the UK100KGP cohort demonstrating coloboma with or without microphthalmia. Inheritance is autosomal dominant with incomplete penetrance and variable expressivity. **B.** Chorioretinal coloboma found in individual A1; *forme fruste* on the right eye (arrow). **C**. Iris coloboma and chorioretinal coloboma found in individual C1. +, individual with variant; -, individual without variant. See Supplementary Table S5 for variant information. **Pedigrees of the Probands and clinical images of eyes are available upon request form the corresponding author.**

**Supplementary Fig. 3:**
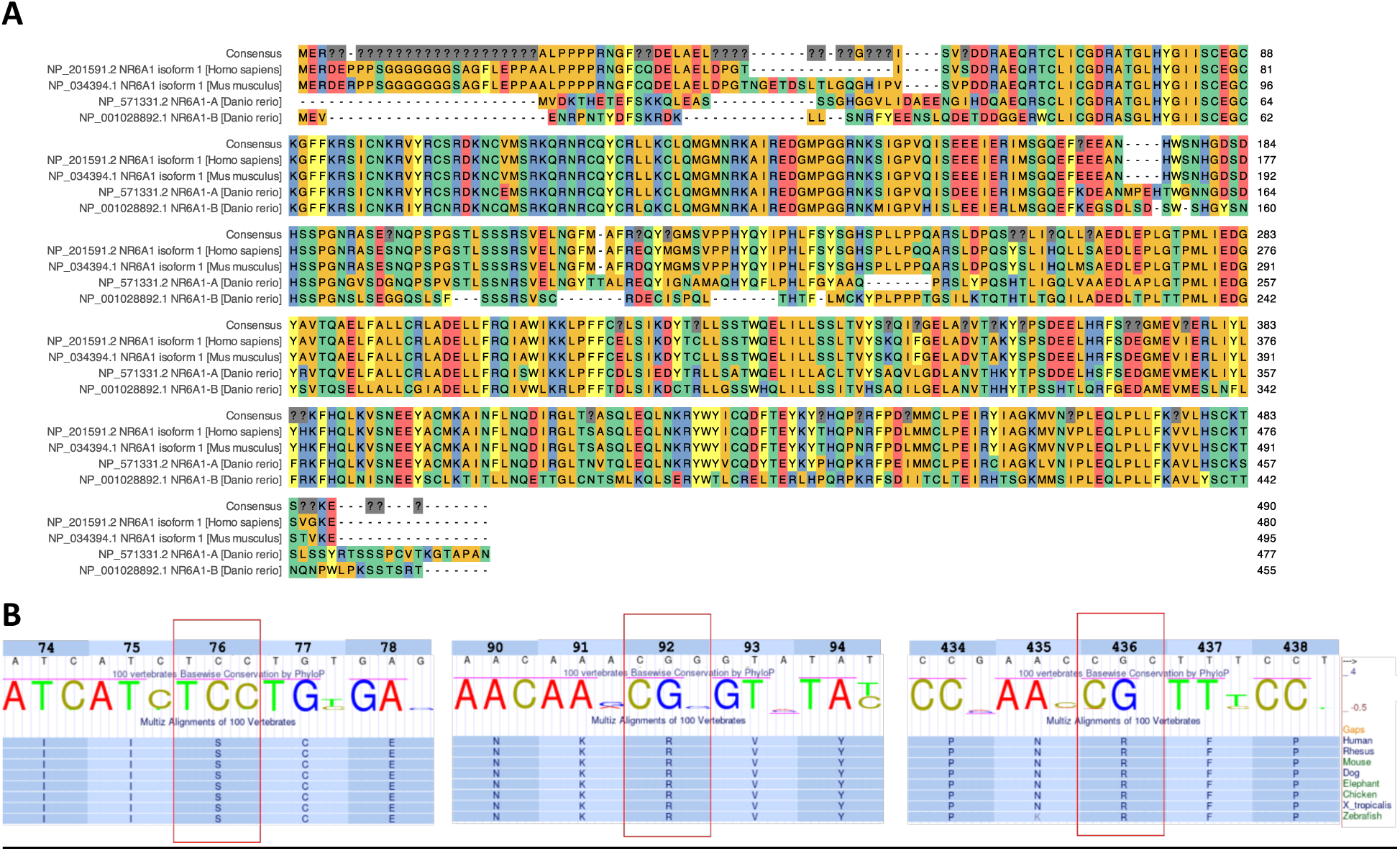
NR6A1 conservation. **A.** Multiple species alignment (MSA) of the canonical transcript of human, mouse, and zebrafish *NR6A1*/*Nr6a1*/*nr6a1* (a and b paralogs) showing a high degree of conservation. **B.** Conservation of Ser76, Arg92 and Arg436 residues during vertebrate evolution. The images for 100 vertebrates Basewise Conservation by PhyloP and Multiz Alignments of 100 Vertebrates were adapted from the UCSC genome browser. Human NR6A1 cDNA sequence and amino acid numbers are listed on the top.

**Supplementary Fig. 4:**
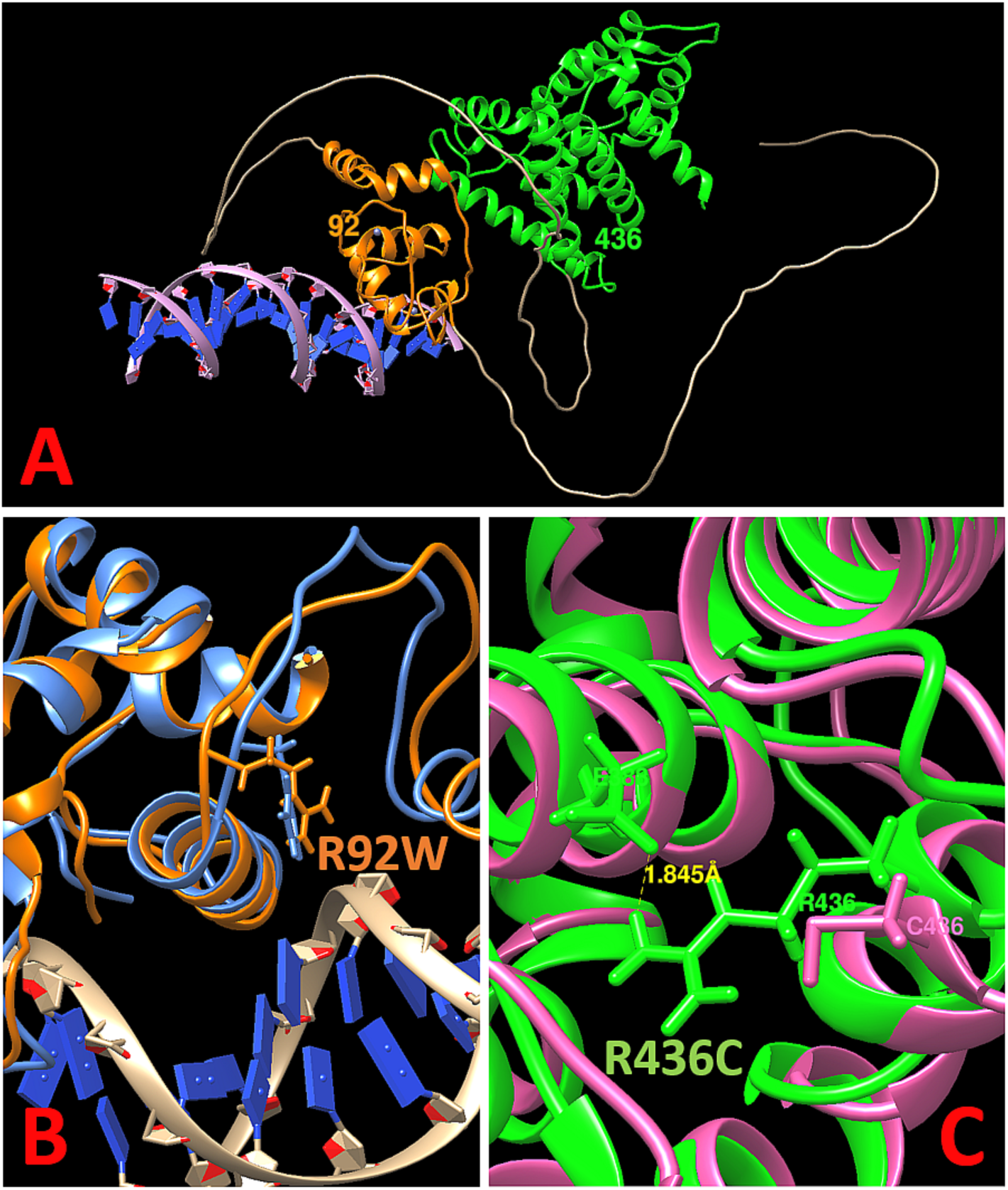
A. *In silico* molecular modeling of NR6A1 with DNA. (lilac helix with blue base pairs). The predicted DNA-binding domain and ligand-binding domain are shown in orange and green, respectively. The positions of R92W and R436 are noted. **B**. The R92W variant changes a positively charged Arg to a hydrophobic Trp and is expected to disrupt interaction with the negatively charged DNA helix. **C.** The R436C variant is predicted to disrupt a hydrogen bond between R436 and E388 and substitute a Cys residue that could form abnormal disulfide bridges within the protein.

**Supplementary Fig. 5:**
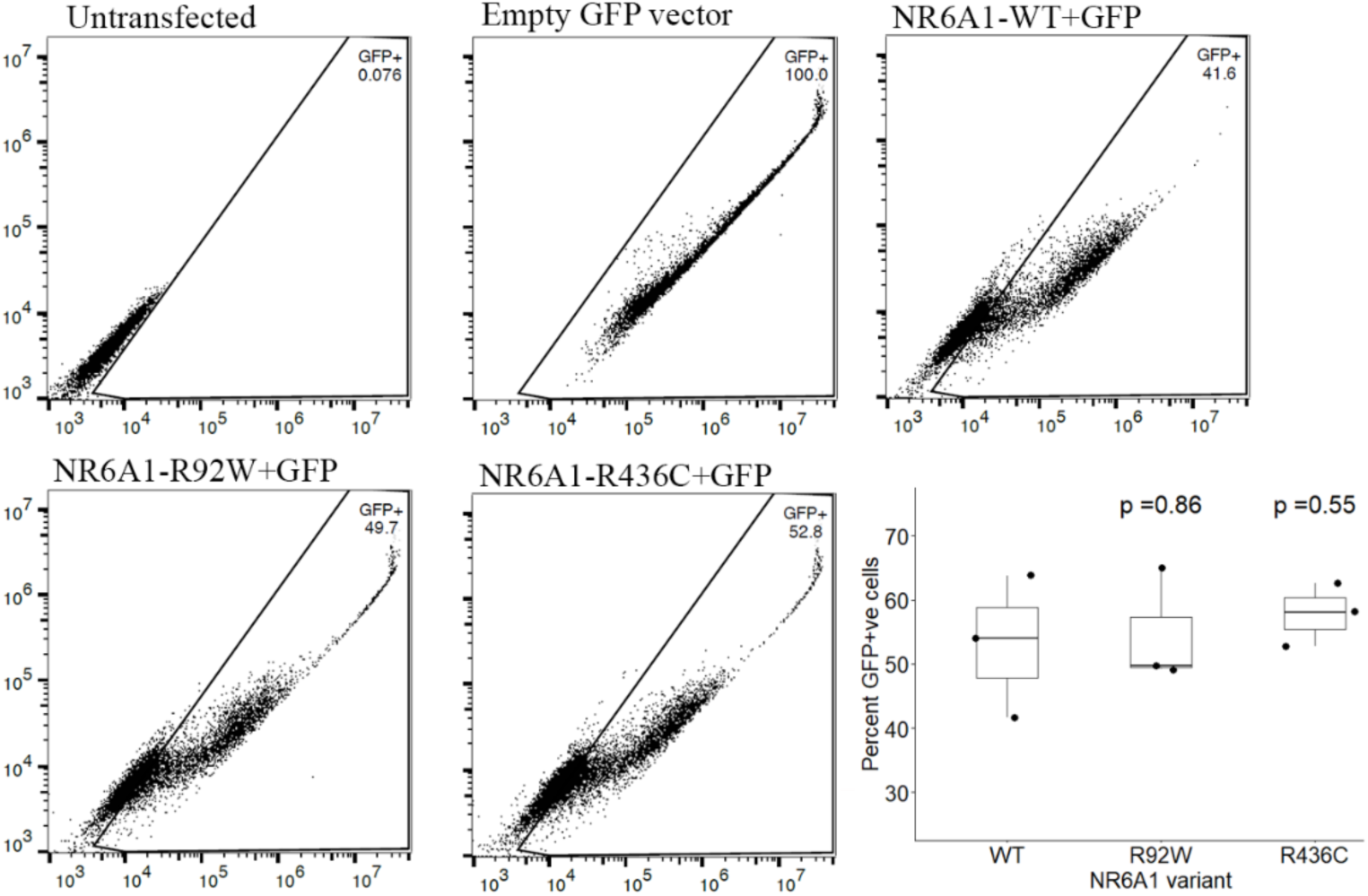
Flow cytometry of wild-type (WT) and mutant (R92W, R436C) forms of NR6A1 demonstrating comparable transfection efficiencies.

**Supplementary Fig. 6:**
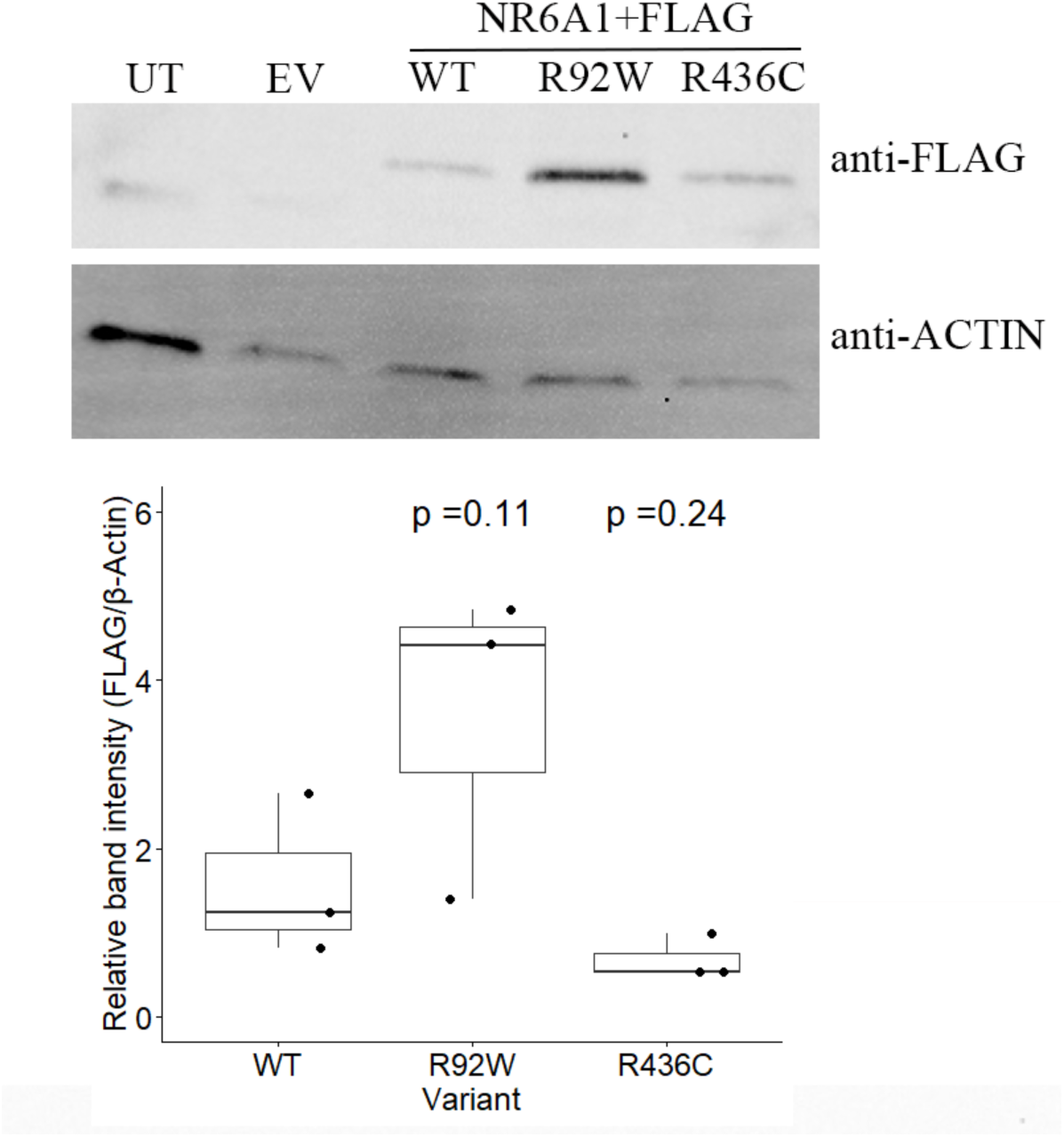
Western blot of HEK293 cells transfected with wild-type (WT) or mutant (R92W, R436W) forms of *NR6A1*, demonstrating comparable transfection efficiencies.

**Supplementary Fig. 7:**
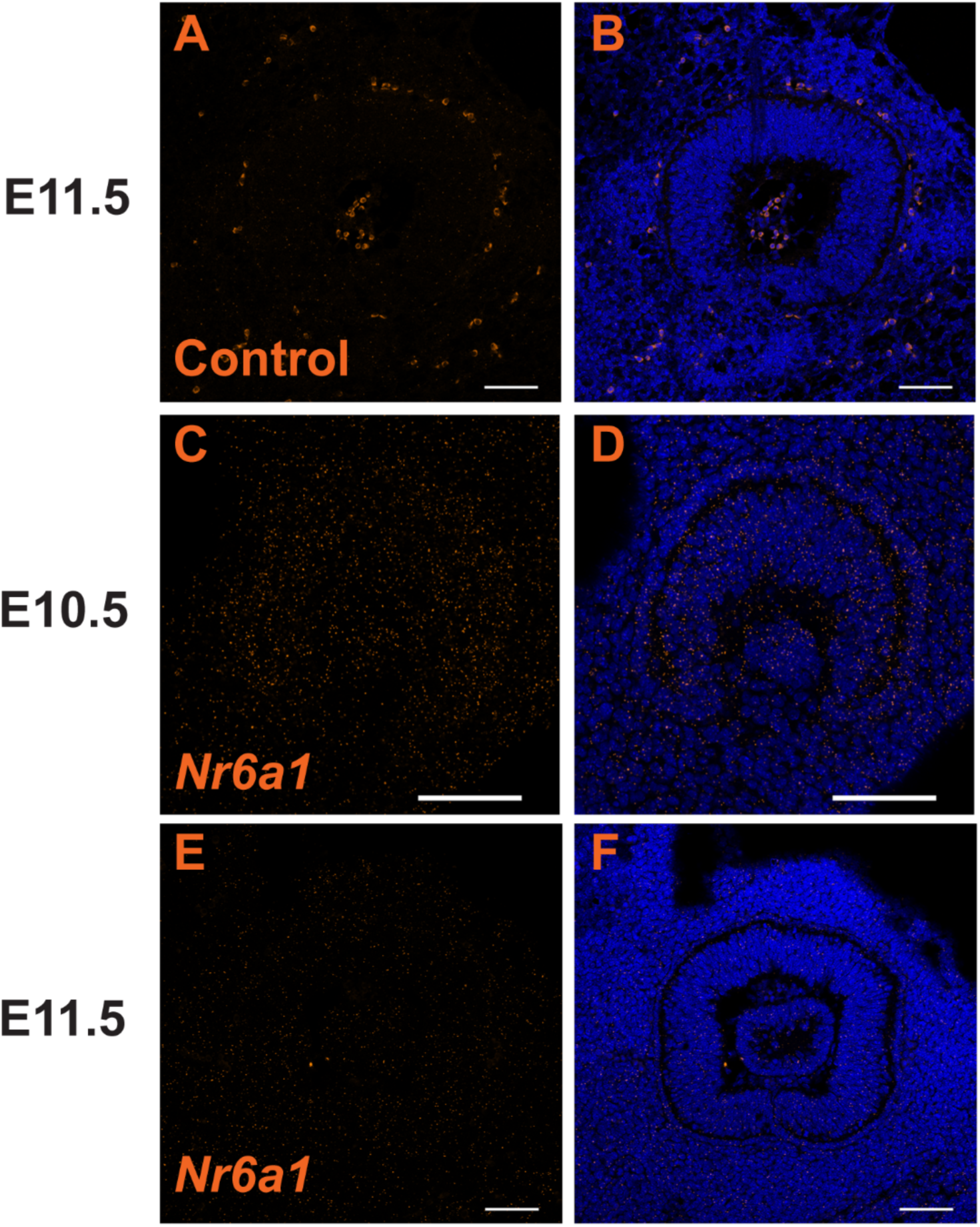
Expression of *Nr6a1* in sagittal sections of mouse embryonic eye before (E10.5) and during (E11.5) optic fissure closure. Low level expression throughout the tissue at E10.5 (C,D) becomes significantly downregulated by E11.5. (E,F). Expression shown with and without DAPI counterstain along with control samples (A,B). Scale bar = 100 μm.

**Supplementary Fig. 8:**
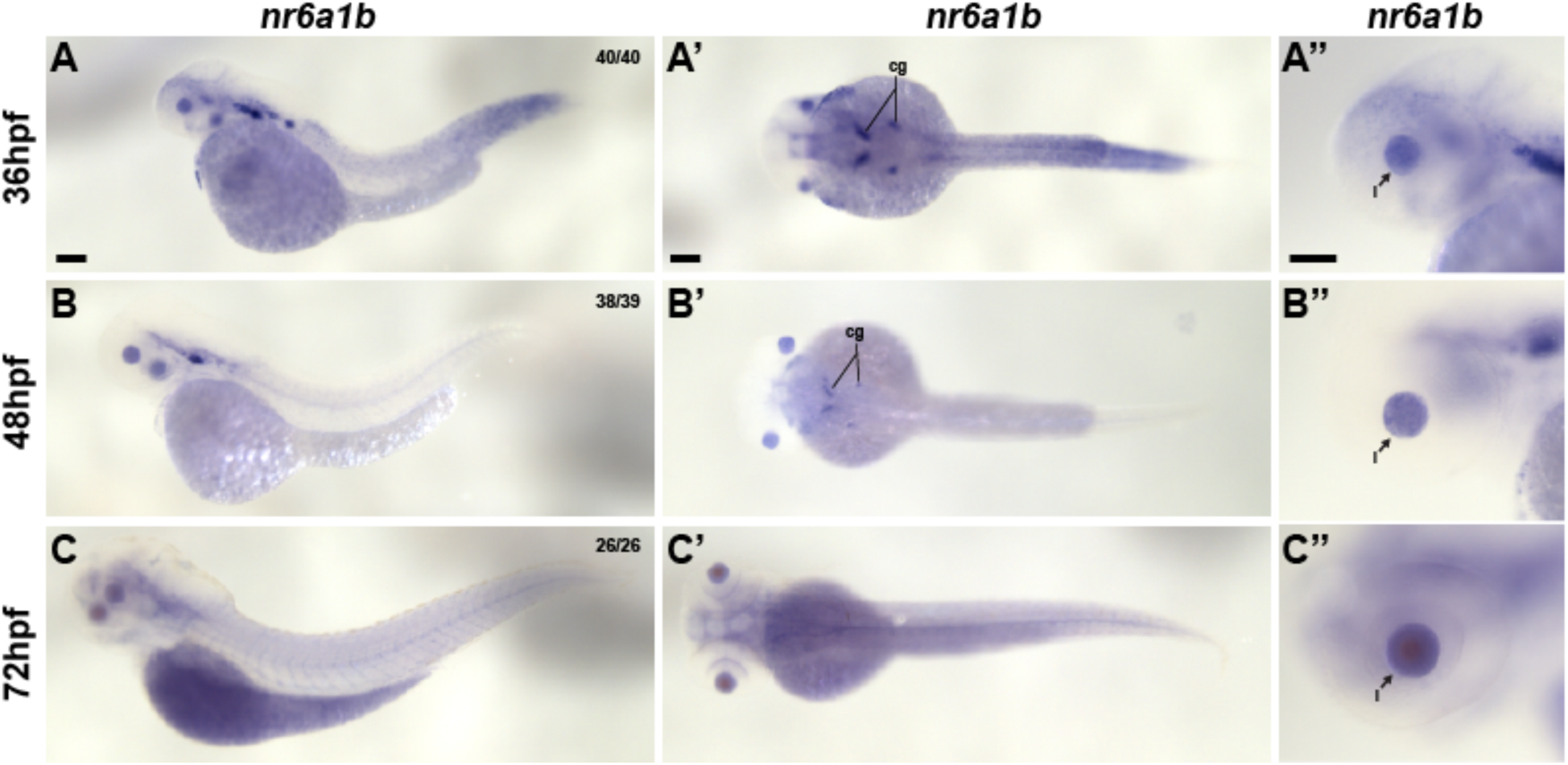
*nr6a1b* mRNA expression during later developmental stages (36 hpf (A-A’’), 48 hpf (B-B’’), and 72 hpf (C-C’’)) of zebrafish development. Note prominent expression in the developing lens (arrow) at all time points and decreased expression in the somites and neural tube, compared to earlier time points. cg-cranial ganglia, l-lens. Scale bar = 100 µM.

**Supplementary Fig. 9:**
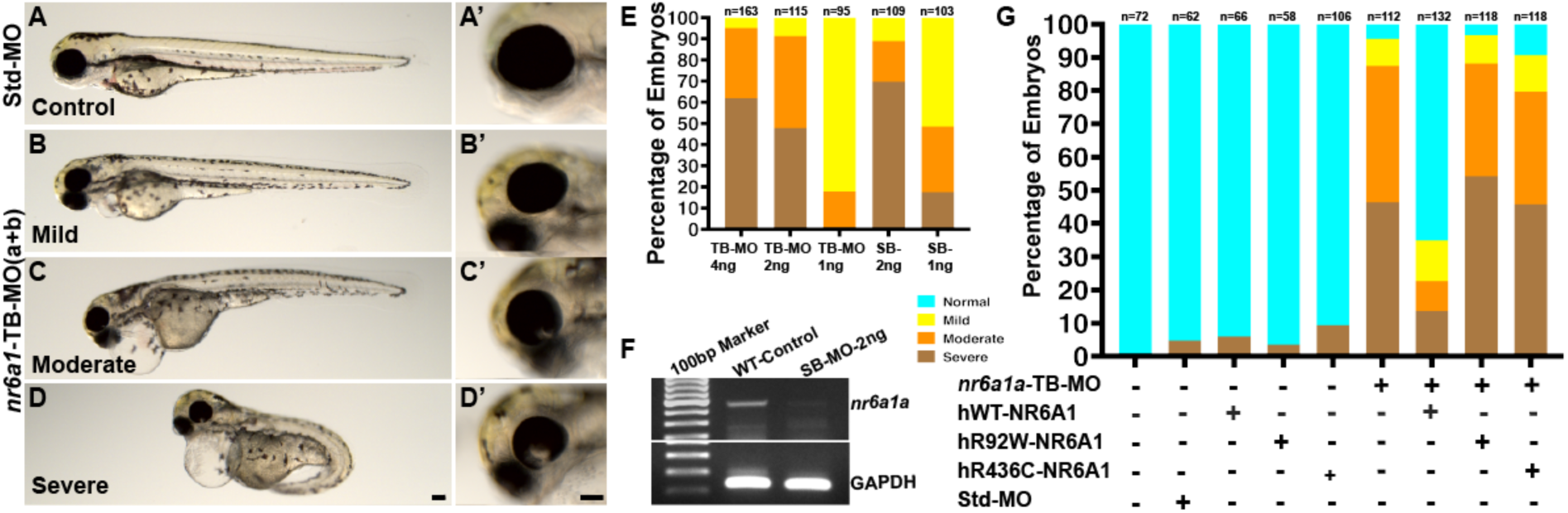
Morpholino (MO) mediated knockdown of the *nr6a1a* paralog with either translation blocking (TB-MO) or splice-blocking (SB-MO) results in a spectrum of phenotypes including colobomatous microphthalmia, shortened/bent body axis and heart edema (A-D, with higher magnification of the eye in the corresponding A’-D’ panel). 2ng of either TB-MO or SB-MO was sufficient to cause 80-90% of embryos to have a moderate or severe phenotype (E). Efficient blocking of the splicing by the SB-MO was confirmed with reverse-transcriptase PCR (F). Although 100pg human wild-type (hWT) mRNA resulted in dramatic phenotypic rescue of the TB-MO (>60% with a normal phenotype), rescue with either hR92W- or hR436W-mRNA was considerably less effective (F). Scale bar = 100µM.

**Supplementary Fig. 10:**
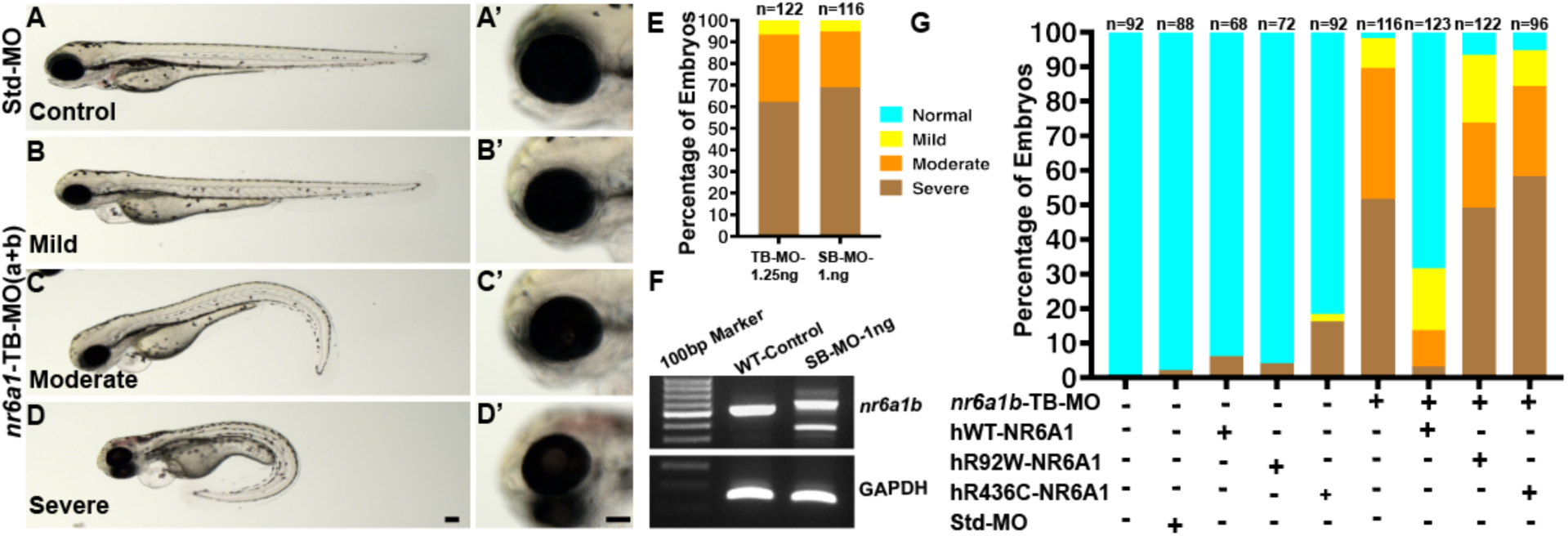
Morpholino mediated (MO) knockdown of the *nr6a1b* paralog with either translation blocking (TB-MO) or splice-blocking (SB-MO) results in a spectrum of phenotypes including microphthalmia, shortened/bent body axis and heart edema (A-D, with higher magnification of the eye in the corresponding A’-D’ panel). 1.25ng and 1ng of TB-MO and SB-MO respectively, was sufficient to cause 80-90% of embryos to have a moderate or severe phenotype (E). Efficient blocking of the splicing by the SB-MO was confirmed with reverse-transcriptase PCR (F). Although 100pg human wild-type (hWT) mRNA resulted in dramatic phenotypic rescue of the TB-MO (>60% with a normal phenotype), rescue with either hR92W- or hR436W-mRNA was considerably less effective (F). Scale bar = 100µM.

**Supplementary Fig. 11:**
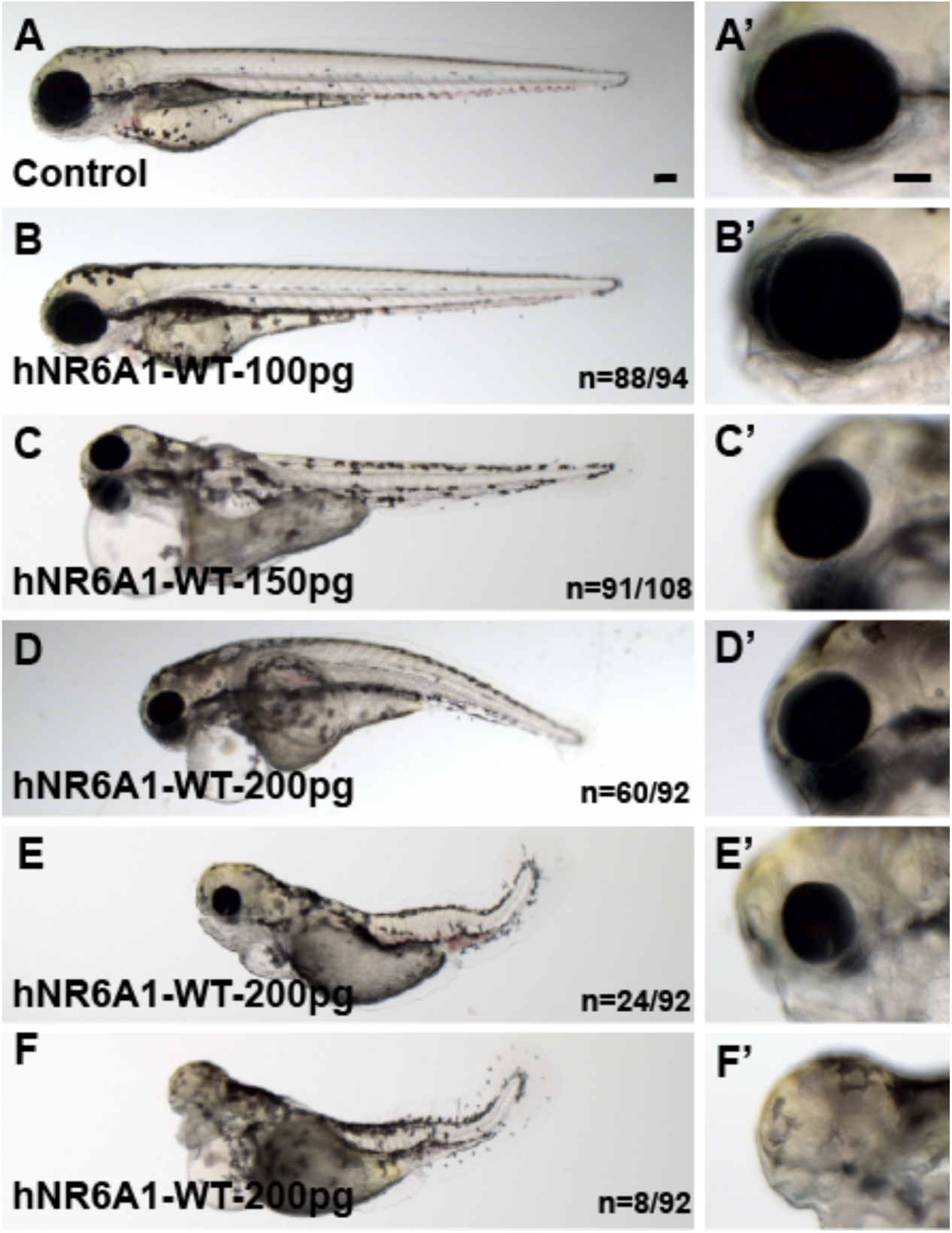
Overexpression of human NR6A1 (*hNR6A1*) in zebrafish: Control and 100pg injected embryos have a normal body with closed OF (A-B’). Embryos injected with 150pg of *hNR6A1* have a shortened but straight body axis, microphthalmia and heart edema (C, C’). A variable phenotype is observed with 200pg of *hNR6A1* RNA, including a bent body axis, heart edema and colobomatous microphthalmia (D, D’) The remaining embryos have a curved body axis with somites losing their chevron shape, microphthalmia with coloboma, and in some cases, lack of eyes and heart edema (E-F’). WT = wild type. Scale bar = 100µM.

**Supplementary Fig. 12:**
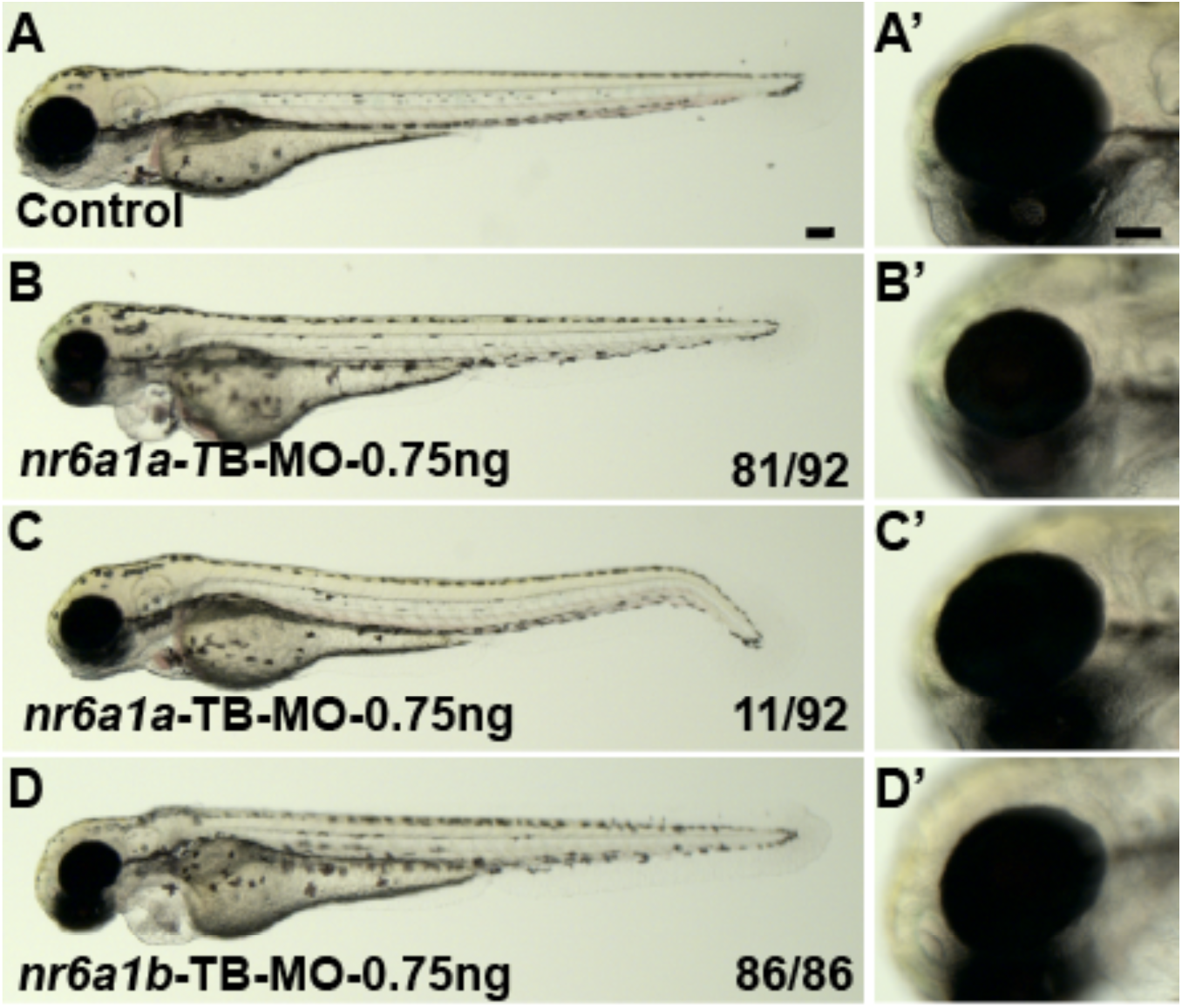
Effect of individual TB-MO for *nr6a1a* and *nr6a1b* at 0.75ng. Control embryos have a straight body and a closed OF (A, A’). Both *nr6a1a* and *nr6a1b* TB morphants at 0.75ng dosage, had straight bodies and microphthalmia, except a few embryos from the *nr6a1a* group which had a slightly curved tail (B-D’). Scale bar = 100µM.

**Supplementary Fig. 13:**
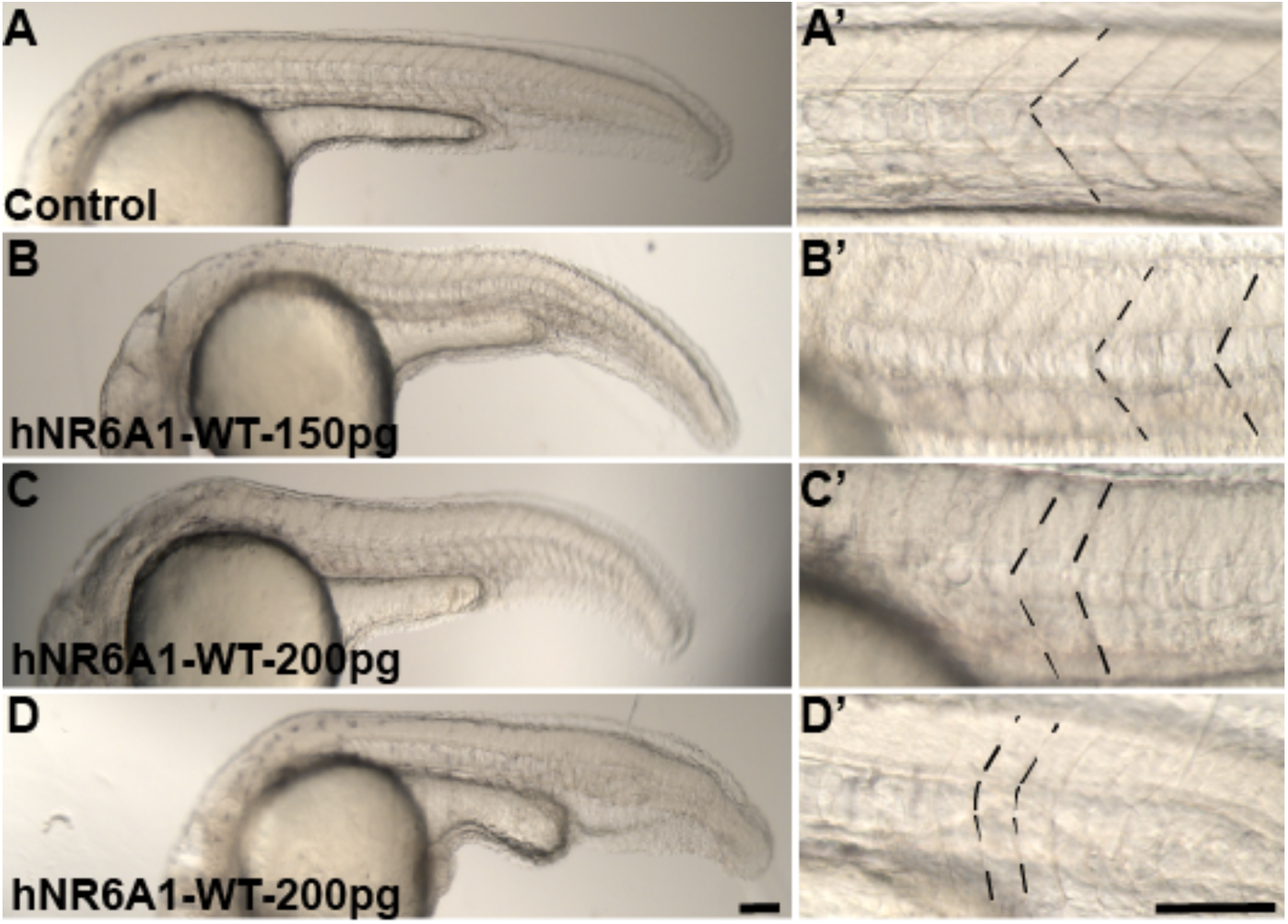
Phenotype of somites in zebrafish embryos over expressed with human NR6A1 (*hNR6A1)*: Control embryos have chevron shaped somites (A, A’), embryos injected with 150pg and 200pf of *hNR6A1* lose the shape of their somites at varying degrees (B-D”). Scale bar = 100µM.

## Notes

### Competing Interest Statement

The authors have declared no competing interest.

### Author Declarations

Institutional Review Board of National Institutes of Health gave ethical approval for this work.

